# Branched chain amino acid metabolism and microbiome in adolescents with obesity during weight loss therapy

**DOI:** 10.1101/2025.02.03.25321363

**Authors:** Jessica R. McCann, Chengxin Yang, Nathan A. Bihlmeyer, Runshi Tang, Tracy Truong, Jie An, Jayanth Jawahar, Olga Ilkayeva, Michael J. Muehlbauer, Zhengzheng Hu, Holly K. Dressman, Lisa Poppe, Joshua A. Granek, Lawrence A. David, Pixu Shi, Pinar Gumus Balikcioglu, Svati H. Shah, Sarah C. Armstrong, Christopher B. Newgard, Patrick C. Seed, John F. Rawls

## Abstract

**BACKGROUND:** Obesity and weight loss in adults have been associated with distinct metabolome and gut microbiome features, but the extent to which those associations apply to adolescent stages remain unclear.

**METHODS:** The Pediatric Obesity Microbiome and Metabolism Study (POMMS) enrolled 220 adolescents aged 10-18 with severe obesity (OB) and 67 healthy weight controls (HWC). Blood, stool, and clinical measures were collected at baseline and after a 6-month obesity intervention for the OB group. Metabolomic profiling in serum using targeted quantitative mass spectrometry and microbiome profiling in stool were performed, and those features were assessed for associations with BMI, insulin resistance, and inflammation. Fecal microbiome transplants were performed on germ-free mice using samples from both groups to assess effects on weight gain and metabolic pathways.

**RESULTS:** Adolescents with OB exhibited higher serum branched-chain amino acid (BCAA) but lower ketoacid metabolite (BCKA) levels compared with HWC. This pattern was sex- and age-dependent, unlike adults with OB, who show elevated levels of both. Longitudinal analysis identified metabolic and microbial features correlated with changes in health measures during the intervention. The fecal microbiomes of adolescents with OB and HWC had similar diversity but differed in membership and functional potential. FMT from both OB and HWC donors had similar effects on mouse body weight, but specific taxa were linked to weight gain in FMT recipients.

**CONCLUSION:** Adolescents with OB have unique metabolomic adaptations and microbiome signatures compared to their HWC counterparts and adults with OB.

**TRIAL REGISTRATION:** ClinicalTrials.gov Identifier: NCT03139877 (Observational Study) and NCT02959034 (Repository)

**FUNDING SOURCES:** American Heart Association Grants: 17SFRN33670990, 20PRE35180195

National Institute of Diabetes and Digestive and Kidney Diseases Grant: R24-DK110492

## INTRODUCTION

Current knowledge about the physiology of obesity and treatment response is dominated by studies conducted in adults. Since the 1960s, studies have established that elevation of circulating branched-chain amino acids (BCAA: valine, leucine, and isoleucine) and various products of their catabolism are associated with adult obesity, metabolic syndrome, and future development of T2D (1–3). The ketoacid metabolites of BCAA (BCKA) have been measured in fewer studies but are also reported to be elevated in adults with obesity (4, 5). Adults who lose weight via dietary and exercise interventions with or without bariatric surgery experience a reduction in BCAA and BCKA levels (6–8). BCAA-deficient diets in rodents confirm a causal relationship with obesity and T2D (9, 10). Circulating glutamate, a product of BCAA metabolism, is also associated with both insulin resistance and obesity in adults (11). Finally, some short and medium chain acylcarnitines, such as the C3 and C5 species generated from BCAA metabolism, are consistently found in a metabolite principal component associated with adult T2D and insulin resistance (2, 12). Together, these results establish that adults with obesity have a metabolic profile consisting of elevated BCAA, BCKA, and associated acylcarnitines and amino acids.

Adolescence is a critical window during which time successful treatment of obesity dramatically reduces the risk for adult obesity, T2D, and cardiovascular mortality, and significant reduction of childhood obesity by late adolescence reduces risk to levels similar to children who never experienced obesity (13, 14). A similar magnitude of weight loss in adults with obesity does not lead to the same degree of risk reduction, suggesting that obesity in earlier life may be driven by physiological mechanisms that are still reversible before adulthood. Yet, little is known about how the metabolic pathways of obesity differ between adolescence and adulthood.

Limited prior work has indicated that obesity in youth aged 4-19 years is characterized by increased BCAA and acylcarnitine catabolism and changes in nucleotides, lysolipids, and inflammatory markers also seen in adults with obesity, as well as reduced fatty acid catabolism that may be unique to obesity in children (15). Recent work comparing male and female adolescents with obesity in the age range of 12-18 years who underwent a 6-month lifestyle intervention program suggests a sex difference in the baseline level of circulating BCKA, with males having higher levels than females despite similar BMI profiles (16). In addition, a recent study showed an elevation in urinary BCAA and BCKA levels in female adolescent subjects with type 2 diabetes compared to obese, non-diabetic, or lean controls, accompanied by diversion of tryptophan metabolism from the serotonin to the kynurenine pathway (17). These metabolic indices may be related to the higher risk of T2D in obese adolescent females than in adolescent males or relatively higher rates of anabolic metabolism in males. However, the similarities and differences in obesity physiology between adolescents and adults remain unclear, especially with respect to sex and age.

Several studies in adults have reported gut microbiome alterations associated with obesity and related diseases (reviewed in (18)). Fecal microbiome transplant (FMT) from adult humans with and without obesity to germ-free (GF) mouse recipients has indicated that the microbiome can causally contribute to obesity and associated metabolic phenotypes. GF mice receiving FMT from adult donors with obesity gain more weight, gain more fat mass, and have distinct microbiota-linked changes to serum metabolite profiles than those receiving FMT from a healthy weight adult donor (19–22). However, it remains unknown if the microbiome causally contributes to obesity physiology during earlier adolescent stages. Based on the lack of gut microbial diversity differences we observed in our preliminary cohort analysis (23) we hypothesized that adolescent obesity would not be directly correlated with gut dysbiosis to the same extent as observed in adults.

We established the Pediatric Obesity Microbiome and Metabolism Study (POMMS) to test the hypothesis that adolescents with obesity have metabolic and microbiome characteristics at baseline and following weight loss intervention that distinguish them from adults with obesity (23). Our study cohort was drawn from the Duke Children’s Healthy Lifestyles program, which offers multi-component behavioral, pharmacotherapeutic, and surgical treatment options as per the current American Academy of Pediatrics guidelines for youth aged 23 and younger with obesity (24). By studying the microbiome and metabolome in this diverse population, we sought to identify new prognostic and therapeutic targets for greater obesity-associated disease risk reduction and reversibility in the most affected populations. Further, adolescents tend to be treatment naïve. Thus, their clinical and metabolic measures are less complicated by drug effects. The study design, data collection, and demographics of the POMMS cohort were previously described (23). Here, we report our findings from the complete metabolome and microbiome datasets at baseline and after intervention.

## RESULTS

### Baseline characteristics

The POMMS clinical cohort was recruited upon referral to the Duke Healthy Lifestyles Clinic in Durham, North Carolina, USA. Details concerning recruitment, treatment plans, and demographics of our study cohort have been previously reported (23) and are summarized in Figure 1A and Supplemental Table 1. Participants with obesity (OB, n= 220) and healthy weight controls (HWC, n= 67) were well-matched according to self-reported race and ethnicity. However, the OB group was significantly more female-identifying and younger than the HWC group (Supplemental Table 1). CRP, HbA1c, ALT, LDL cholesterol, triglyceride, insulin, systolic and diastolic blood pressures, and triglyceride/HDL ratio were higher in the OB group compared with the HWC group (Table 1). HDL cholesterol levels were lower in the OB group as compared with the HWC group. Fasting glycerol levels were also significantly higher in the OB group using a targeted lab analysis (Supplemental Table 1.1), suggesting higher levels of lipolysis in adolescents with OB as compared with HWC (25). However, these two groups had no significant difference in fasting glucose levels, suggesting that even though several metrics associated with OB in adults were significantly higher in the OB cohort when compared to HWC, high BMI alone is not enough to significantly confer a difference in fasting glucose commonly observed in adults with obesity (26).

**Figure 1.**
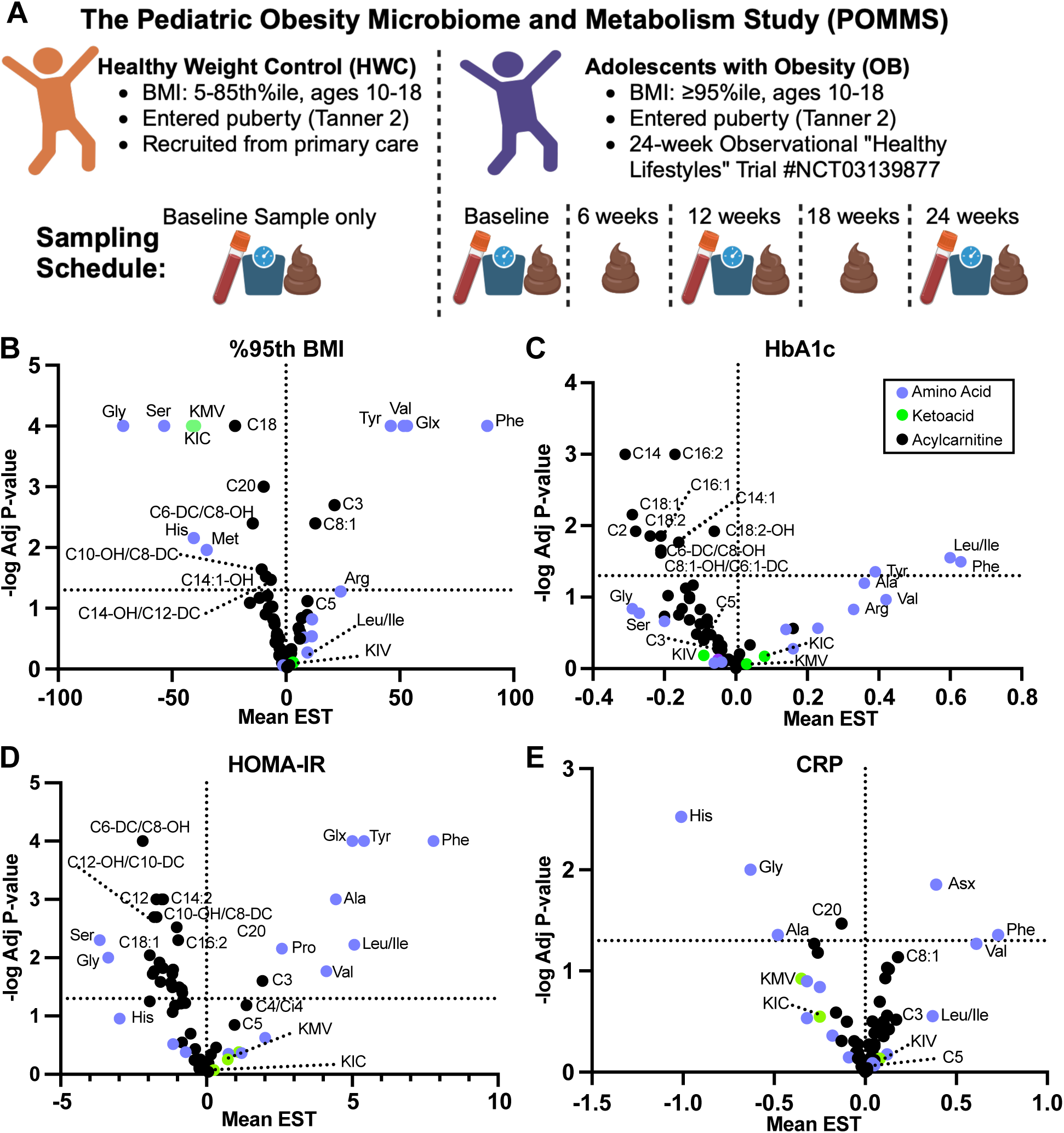
Significant metabolic changes are associated with clinical measures of obesity in adolescents. **(A)** POMMS study overview depicting the inclusion criteria, sampling schedule for both the healthy weight control (HWC) and participants with obesity (OB). **(B-D)** Linear regression analysis adjusted for age, race and sex of participant. See Statistical Analysis description in Methods for details. **(B)** Targeted serum metabolites associated with %95th BMI, **(C)** Hemoglobin A1c (HbA1c), (D) Homeostatic Insulin Resistance score (HOMA-IR), at baseline, or **(E)** C-Reactive Protein (CRP), at baseline. For B-E, metabolites above the dotted line are considered significantly associated with the noted clinical measure. Higher levels of metabolites in the right upper quadrant are significantly associated with higher levels of the clinical measure. In comparison, higher levels of metabolites in the left upper quadrant are significantly associated with lower levels of the clinical measure. %95th BMI is defined as the percent of the BMI above the 95th percentile. Diagram in part **(A)** was created with Biorender.

**Table 1.** Baseline clinical lab values in OB versus HWC groups.

|  | **HWC** | **OB** | **Total** | **OB/HWC**<br>**geometric**<br>**means (95%**<br>**CI)** | **P-**<br>**value\*** |
| --- | --- | --- | --- | --- | --- |
|  | **N=67** | **N=220** | **N=287** |  |  |
| **ALT** |  |  |  | 1.65 (1.43,<br>1.89) | <0.001 |
| Mean (SD) | 14.6 (6.7) | 24.7 (31.8) | 22.9 (29.1) |  |  |
| Median (Q1,<br>Q3) | 13.0 (11.0, 15.2) | 20.0 (15.0,<br>25.0) | 18.0 (14.0, 24.0) |  |  |
| (Min, Max) | (6.0, 44.0) | (10.0, 456.0) | (6.0, 456.0) |  |  |
| Missing | 19 (28.4%) | 4 (1.8%) | 23 (8.0%) |  |  |
| **HbA1c** |  |  |  | 1.04 (1.01,<br>1.07) | 0.007 |
| Mean (SD) | 5.3 (0.3) | 5.6 (0.6) | 5.5 (0.5) |  |  |
| Median (Q1,<br>Q3) | 5.3 (5.2, 5.5) | 5.5 (5.3, 5.7) | 5.5 (5.2, 5.7) |  |  |
| (Min, Max) | (4.4, 6.0) | (4.5, 11.7) | (4.4, 11.7) |  |  |
| Missing | 20 (29.9%) | 10 (4.5%) | 30 (10.5%) |  |  |
| **LDL** |  |  |  | 1.16 (1.05,<br>1.28) | 0.003 |
| Mean (SD) | 88.0 (27.3) | 97.0 (25.7) | 95.3 (26.2) |  |  |
| Median (Q1,<br>Q3) | 82.0 (72.0, 101.0) | 94.0 (80.2,<br>111.0) | 92.0 (78.0,<br>110.0) |  |  |
| (Min, Max) | (38.0, 168.0) | (9.0, 195.0) | (9.0, 195.0) |  |  |
| Missing | 18 (26.9%) | 6 (2.7%) | 24 (8.4%) |  |  |
| **HDL** |  |  |  | 0.84 (0.78,<br>0.90) | <0.001 |
| Mean (SD) | 53.7 (12.3) | 45.9 (10.1) | 47.3 (10.9) |  |  |
| Median (Q1,<br>Q3) | 51.0 (45.0, 59.0) | 45.0 (39.0,<br>52.0) | 46.0 (40.0, 53.0) |  |  |
| (Min, Max) | (36.0, 100.0) | (27.0, 77.0) | (27.0, 100.0) |  |  |
| Missing | 18 (26.9%) | 5 (2.3%) | 23 (8.0%) |  |  |
| **Triglyceride** |  |  |  | 1.31 (1.14,<br>1.52) | <0.001 |
| Mean (SD) | 84.3 (43.6) | 111.6 (56.7) | 106.5 (55.4) |  |  |
| Median (Q1,<br>Q3) | 72.0 (52.0, 97.0) | 97.0 (73.0,<br>139.0) | 91.0 (69.0,<br>133.8) |  |  |
| (Min, Max) | (29.0, 223.0) | (22.0, 404.0) | (22.0, 404.0) |  |  |
| Missing | 18 (26.9%) | 5 (2.3%) | 23 (8.0%) |  |  |
| **TG/HDL ratio** |  |  |  | 1.57 (1.32,<br>1.87) | <0.001 |
| Mean (SD) | 1.7 (1.1) | 2.6 (1.7) | 2.5 (1.6) |  |  |
| Median (Q1, Q3) | 1.3 (0.9, 2.1) | 2.1 (1.5, 3.2) | 2.0 (1.3, 3.1) |  |  |
| (Min, Max) | (0.5, 5.9) | (0.4, 11.9) | (0.4, 11.9) |  |  |
| Missing | 18 (26.9%) | 5 (2.3%) | 23 (8.0%) |  |  |
| **C-reactive protein** |  |  |  | 6.58<br>(4.34, 9.97) | <0.001 |
| Mean (SD) | 0.2 (0.7) | 0.6 (0.9) | 0.5 (0.9) |  |  |
| Median (Q1, Q3) | 0.0 (0.0, 0.1) | 0.3 (0.1, 0.6) | 0.2 (0.1, 0.5) |  |  |
| (Min, Max) | (0.0, 4.4) | (0.0, 6.9) | (0.0, 6.9) |  |  |
| Missing | 20 (29.9%) | 7 (3.2%) | 27 (9.4%) |  |  |
| **Insulin** |  |  |  | 1.82<br>(1.46, 2.25) | <0.001 |
| Mean (SD) | 11.3 (10.1) | 19.0 (17.3) | 17.5 (16.5) |  |  |
| Median (Q1, Q3) | 7.7 (5.6, 12.2) | 15.4 (10.6, 21.8) | 13.9 (8.6, 20.5) |  |  |
| (Min, Max) | (3.4, 52.0) | (3.2, 150.0) | (3.2, 150.0) |  |  |
| Missing | 20 (29.9%) | 25 (11.4%) | 45 (15.7%) |  |  |
| **Glucose** |  |  |  | 1.03<br>(0.99, 1.07) | 0.117 |
| Mean (SD) | 86.7 (9.0) | 88.8 (15.3) | 88.4 (14.3) |  |  |
| Median (Q1, Q3) | 87.0 (83.0, 93.0) | 87.0 (83.0, 92.0) | 87.0 (83.0, 92.0) |  |  |
| (Min, Max) | (64.0, 108.0) | (69.0, 264.0) | (64.0, 264.0) |  |  |
| Missing | 18 (26.9%) | 5 (2.3%) | 23 (8.0%) |  |  |
| **Systolic Blood Pressure** |  |  |  | 1.04<br>(1.02, 1.07) | <0.001 |
| Mean (SD) | 110.9 (10.9) | 113.0 (9.7) | 112.5 (10.0) |  |  |
| Median (Q1, Q3) | 110.0 (102.0, 120.0) | 112.0 (108.0, 120.0) | 112.0 (106.0, 120.0) |  |  |
| (Min, Max) | (90.0, 151.0) | (80.0, 140.0) | (80.0, 151.0) |  |  |
| Missing | 0 (0%) | 1 (0.5%) | 1 (0.3%) |  |  |
| **Diastolic Blood Pressure** |  |  |  |  |  |
| Mean (SD) | 65.7 (8.1) | 68.9 (7.7) | 68.1 (7.9) | 1.07<br>(1.04, 1.11) | <0.001 |
| Median (Q1, Q3) | 64.0 (60.0, 70.5) | 70.0 (62.0, 73.5) | 68.0 (62.0, 72.0) |  |  |
| (Min, Max) | (48.0, 82.0) | (50.0, 96.0) | (48.0, 96.0) |  |  |
| Missing | 0 (0%) | 1 (0.5%) | 1 (0.3%) |  |  |

### Serum metabolites reveal distinct adolescent sex- and age-dependent associations with BMI, insulin resistance, and inflammation

In an adjusted analysis of serum metabolites in OB and HWC adolescents at baseline, levels of BCAA valine (Val), the amino acids tyrosine (Tyr), glutamine/glutamate (Glx), and phenylalanine (Phe), and C3 (propionyl, a bioproduct of BCAA catabolism) and C8:1 acylcarnitines were all significantly and positively associated with BMI measured as a percent of the BMI 95^th^ percentile (%95^th^ BMI) (27). In contrast, higher serum levels of the amino acids glycine (Gly), serine (Ser), histidine (His), methionine (Met), and several even chain acylcarnitines were significantly negatively associated with %95^th^ BMI (Figure 1C), as were the ketoacids of BCAAs, leucine (KIC) and isoleucine (KMV). This is in notable contrast to adults in which these ketoacids are typically positively associated with obesity and T2D (4, 7). Several of the metabolites associated with %95^th^ BMI at baseline were also associated with HOMA-IR at baseline, including those negatively associated (Gly, Ser, and acylcarnitines C20 and C6-DC/C8-OH) and positively associated (Val, Tyr, Phe, Glx, C3) with HOMA-IR (Figure 1D). HOMA-IR was also significantly positively associated with levels of leucine/isoleucine (Leu/Ile) and proline (Pro) but was not associated with BCKA. In adults, elevated BMI and HOMA-IR have similarly been associated with higher BCAA, Phe, Tyr, and C3 acylcarnitine (2, 4, 5). However, adult BMI and HOMA-IR are also associated with high levels of BCKA as noted above, but we did not observe this in our adolescent cohort. These data suggest that adolescents with OB have metabolic features like adults with obesity including higher BCAA and associated metabolites, but also display adaptations that differ from adults including lower BCKA in association with high BMI.

In analyses of HbA1c, only the amino acids Leu/Ile, Phe, and Tyr were significantly positively associated with HbA1c (Figure 1C); the acylcarnitines C3-C5, and BCKA KIV were negatively (but not significantly) associated with higher HbA1c, again suggesting metabolites associated with IR and OB in adults are not significantly associated with key clinical measures of metabolic syndrome in our adolescent cohort. It should be noted that HbA1c is a predictive measure for IR in adults, but its use as a measure of metabolic health during adolescence can sometimes be difficult to interpret, as high levels can occur in adolescents with a normal weight and fasting glucose profiles (28).

CRP, a clinical marker of general inflammation, significantly correlated with fewer metabolites either positively (Val, Phe, Asx) or negatively (acylcarnitine C20, His, Gly, and alanine Ala) at baseline (Figure 1E). Some of these significant CRP-metabolite associations were similar in directionality to those with %95^th^ BMI or HOMA-IR (Val, Phe, His, Gly, C20), whilst others were unique to CRP (Asx) or exhibited reversed directionality compared to HOMA-IR (Ala). Whereas only a subset of acylcarnitine species measured here were significantly associated with these three clinical measures, we observed that most acylcarnitines were generally positively correlated with CRP but negatively correlated with HOMA-IR and %95^th^ BMI. Therefore, the general association trend between most serum acylcarnitines and CRP is inverse to that of %95^th^ BMI, HbA1c, and HOMA-IR.

### BCAA metabolism in adolescent obesity is sex and age dependent

Prior work suggests that there is an interaction between obesity status, pubertal status, sex and age that influences BCAA and their related metabolite levels (16). To further assess these metabolic signatures in our adolescent cohort, we analyzed data stratified by OB v HWC status and age on serum metabolite levels to identify factors that influence metabolite levels in our cohort. We found initially that signatures associated with OB v HWC status were driven by metabolite levels in HWC males (Figure 2, Supplemental Table 2). We also found that serum levels of the BCAA, along with several other amino acids and acylcarnitines involved in BCAA metabolism, were significantly varied by OB v HWC status and sex (Figure 2A). Males generally had higher levels of most affected amino acids and their metabolites. When we analyzed raw data and adjusted only for multiple comparisons, males with HWC status had significantly higher levels of Leu/Ile, Glx, and the BCAA metabolite C3 than OB males, as well as the ketoacids of Leu and Ile (KIC and KMV, respectively; Figure 2B-J). Following adjustment for age and race, several of these relationships remained significant, including higher levels of Leu/Ile, C8:1-OH/C6:1DC, C18, C20-OH/C18-DC, Met, His, Orn, and Cit in HWC males compared with males with OB (Supplemental Table 2.1). Because adults with OB tend to have higher levels of BCKA compared to HWC (4–8), we tested if the opposite trends in the adolescent data were still seen in older adolescents by assessing metabolites affected by the interaction of obesity, sex, age, and Tanner (pubertal) stage (29). We found that the amino acids Leu/Ile, His, Orn, Val, Met, Cit, and Phe, and the acylcarnitines C18, C20-OH/C18-DC, C16, C18:1, and C8:1-OH/C6:1DC were significantly affected by the overall interaction between HWC v OB status and sex (Figure 3A, Supplemental Table 2). Second stage testing further delineated the role of status and sex for each indicated metabolite (Supplemental Table 2.1). Based on the interaction of Tanner stage and HWC v OB status, KMV (ketoacid of Ile) was significantly associated with Tanner stages 4 and 5, suggesting that more mature teens had a greater difference in serum levels of this ketoacid (Figure 3B, Supplemental Table 3). To test whether BCKA are associated with sex and obesity at younger ages, we analyzed metabolite data from the Hearts and Parks (HP) Study, which included children enrolled as early as age 5 (30). In these analyses of children aged 5-9 years, we found that the ketoacid KMV was no longer associated with sex in the younger children. Still, the BCAA acylcarnitine metabolite C3 was associated with sex in the older age group after stratified analysis (Figure 3C). These results indicate that obesity-associated differences in BCAA and related metabolite levels are sex-dependent and emerge later in adolescence.

**Figure 2.**
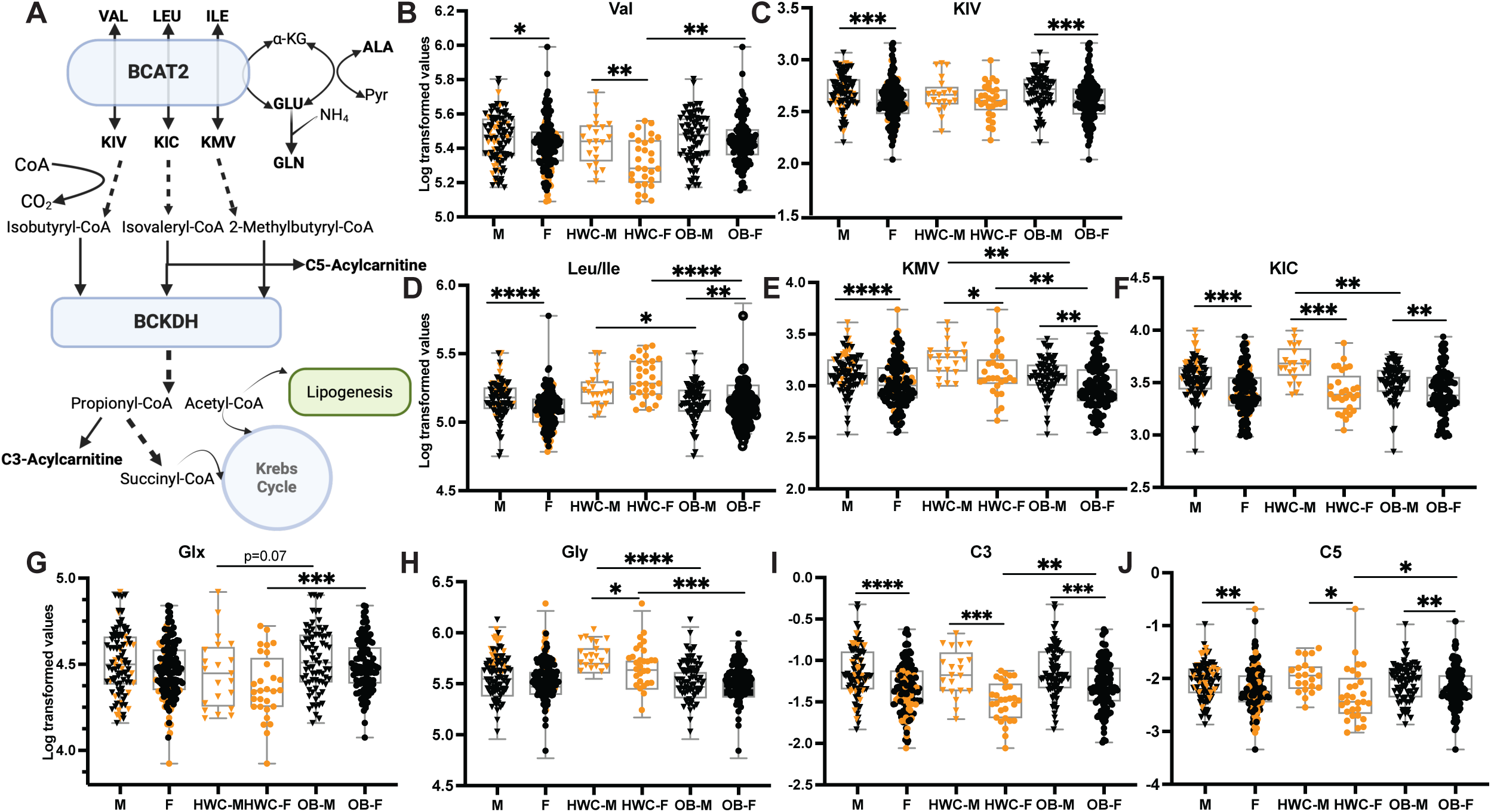
Branched chain amino acids (BCAA) and related metabolite levels are sex dependent, significantly different in HWC v OB cohorts, and unique to adolescents with obesity. **(A)** BCAA related metabolism pathway. Valine (VAL), Leucine (LEU) and Isoleucine (ILE) are reversibly transaminated by the branched chain amino acid transferase (BCAT2) in the mitochondria to their respective ketoacids ketoisovalerate (KIV), ketoisocaproate (KIC) and ketomethylvalerate (KMV). The amino group removed from the BCAA is transferred to α-ketoglutarate (α-KG), yielding glutamate (GLU). The ketoacids can then be irreversibly hydrolyzed by the branched chain ketoacid dehydrogenase (BCKDH) where BCAA derived carbons can then enter the TCA cycle or contribute to lipogenesis. When BCAA are elevated, as in obesity, excess BCAA increases BCAT2 activity which could result in an increased nitrogen load, especially in muscle tissues. This load can be relieved, most likely via coordinated action of serine dehydratase, serine hydroxymethyltransferase, and glycine acyltransferase. Resulting increased metabolites, such as glutamine (GLN) and acylglycine can be secreted from the affected tissues. ALA, alanine; PYR, pyruvate. **(B-J)** Levels of serum amino acids and keto acids associated with BCAA and alanine-pyruvate metabolism in HWC v OB cohorts divided by sex. Box plots represent mean and standard deviation. Statistics were performed on mean metabolite values following log transformation, and P values result from Mann-Whitney tests and are corrected for multiple comparisons. **, P<0.01; ***, P<0.001; ****, P<0.0001.

**Figure 3.**
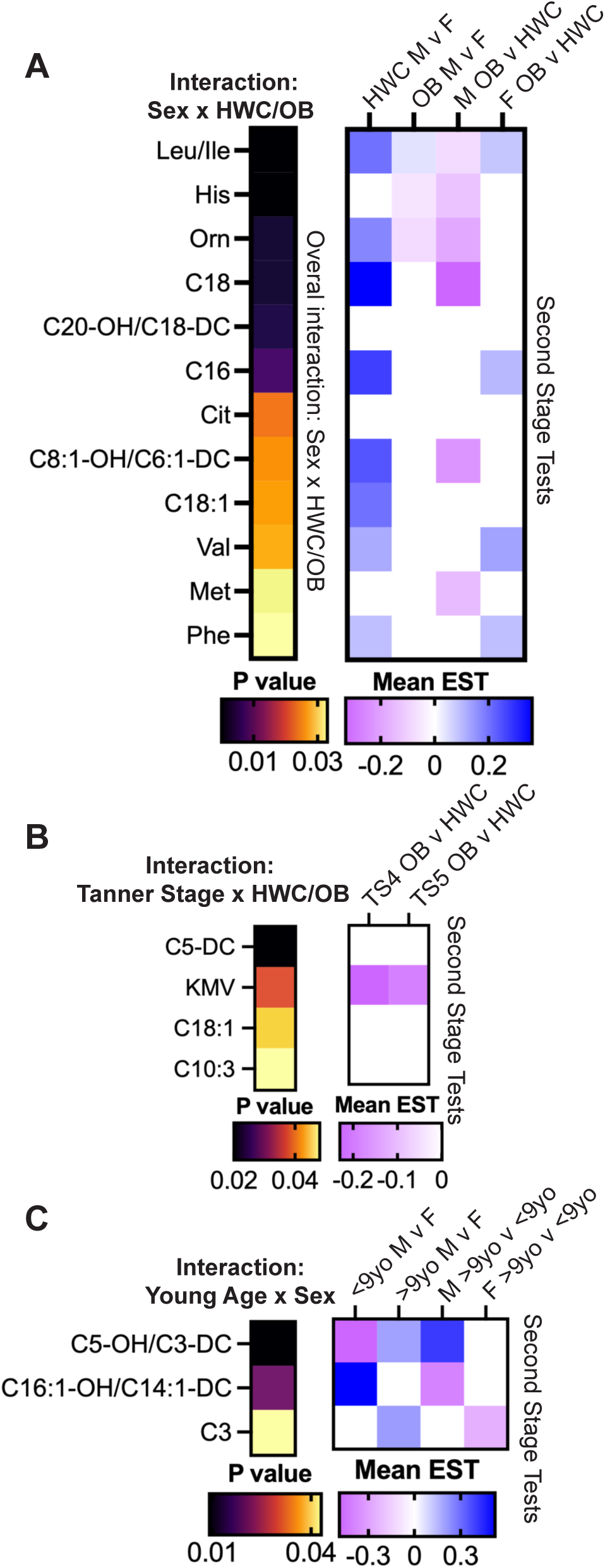
Sex-dependent differences in serum ketoacids are less evident in younger children than in more mature adolescents. Log-transformed levels of serum metabolites tested for significance in an overall interaction and between **(A)** sex and HWC v OB status; **(B)** Tanner (puberty) stage and HWC v OB status; and **(C)** Young age (9 y or younger) versus older age metabolites and sex. Only significant metabolites are shown, P < 0.05, after adjustment for race and multiple comparisons. Complete metabolite results are shown in Supplemental Tables 2, 2.1, 3, and 3.1). See Statistical Analysis methods for details.

### Microbiome alpha and beta diversity were similar between OB and HWC adolescents, but taxa and microbial features were distinct between cohorts

We analyzed 16S rRNA gene amplicon sequence data from 786 fecal samples covering 50 HWC participants at baseline, all 5 timepoints from 57 participants with OB that contributed fecal samples at 0, 1.5, 3, 4.5 and 6 months of the POMMS study, and at least 0 and 6 month timepoints from 140 participants in the OB cohort. As in the interim analysis (23), we observed no significant differences in either alpha or beta diversity based on HWC v OB status at baseline, sex, or age at entry (Supplemental Figure 1A and data not shown). When we tested whether specific bacterial lineages might be associated with %95^th^ BMI at baseline, we found that relative abundance of several family-level taxa were positively correlated with %95^th^ BMI, while the HWC group had significantly higher abundance in members of the Lachnospiraceae and Bacteroiodaceae compared with the OB group (Supplemental Figure 1B).

To gain further insight into these microbial communities, shotgun DNA sequencing data were obtained for 396 participant-derived fecal samples, including quality controls for sequencing fidelity and sampling replication. High quality sequences were obtained for 140 OB at baseline and 6 months, and 50 HWC participants at the baseline time point. Consistent with the 16S rRNA data, alpha and beta diversity did not significantly differ between the OB and HWC cohorts (Supplemental Figure 2), and profiles of the most abundant family- and species-level taxa were generally similar between the two cohorts (Supplemental Figure 3). Further analysis determined, however, that participants from the OB cohort had a significantly higher Firmicutes to Bacteroidetes ratio than HWC (P= 0.001). ALDEx2 (31), ANCOM-BC (32), and corncob (33) analyses were employed to identify differential taxa between the two groups at baseline. The methods detected an increased relative abundance of genera *Ligilactobacillus* and reduced *Alistipes* among the OB participants. Of the *Ligilactobacillus*, the species *L. ruminis* was most prevalent (>90% of samples), and of *Alistipes*, the species *A. finegoldii* was found in most samples (>90%). Notably, *Ligilactobacillus* and *Alistipes* abundance remained positively and negatively correlated, respectively, with %95th BMI (Figure 4A).

**Figure 4.**
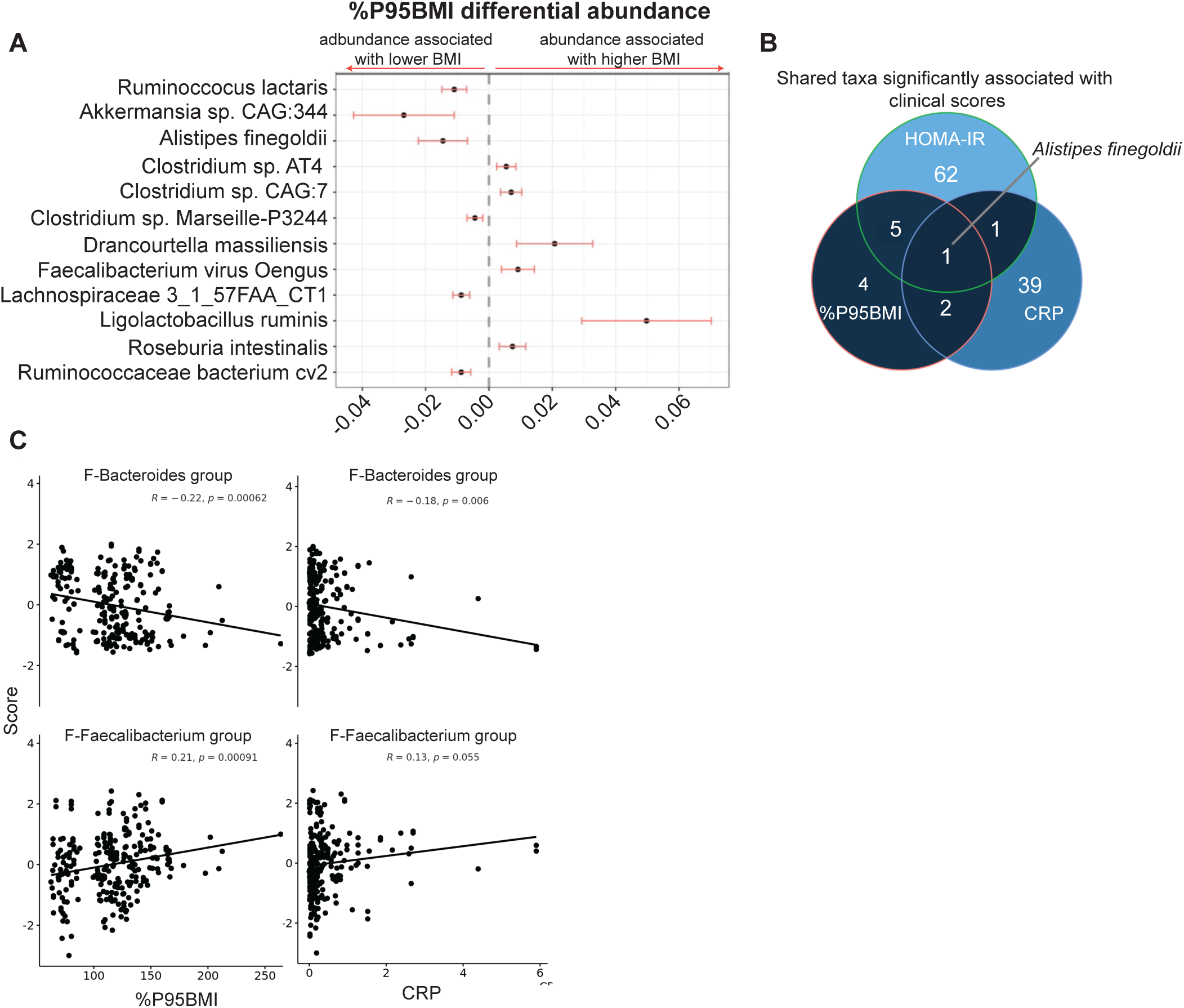
Several gut microbial taxa and predicted enzymatic functions are associated with clinical variables at baseline. **(A)** Abundance of microbial species determined by fecal metagenomic sequencing that were positively or negatively significantly associated with %95th BMI at baseline. **(B)** Venn diagram showing shared and distinct numbers of fecal microbial species associated with %95th BMI, insulin resistance score (HOMA-IR) and C-reactive protein (CRP) levels at baseline. **(C)** Linear regression correlation following dimensionality reduction between co-varying microbial factors and clinical variables at baseline. Factors are named for prominent taxa within each cluster, and the underlying taxa and weights in each cluster is listed in Supplemental Tables 4.1-4.4.

To reduce data dimensionality, taxa and functional capacity of the gut microbiomes were clustered using hierarchical clustering, and the resulting factors were associated with clinical scores at baseline (Figure 4C, Supplemental Figure S4, and Supplemental Tables 4.1-4.3). Though microbial species diversity was similar between the two cohorts at baseline (Supplemental Figure 2), we identified two taxonomic clusters significantly associated with either %95^th^ BMI or CRP as continuous variables (Figure 4C, with factors defined in Supplemental Tables 4.1-4.4). Further clustering of serum metabolomics, KEGG orthology functions, and taxa revealed associations with clinical variables. We found several clusters of covarying taxa, genes, phages, and serum metabolites indicating that there is cross talk between metabolomic status and the microbiome (Supplemental Figure 4, with weights and factors defined in “Supplemental Tables – Factor Weights”). Using a random forest machine learning model, we generated a model that predicts OB v HWC status, which was significantly enriched for pathways involved in microbial amino acid synthesis, two-component signaling, lipopolysaccharide biosynthesis, and bacterial cell membrane components. Notably, several genes required for microbial synthesis of BCAA and tryptophan were independently identified as significant contributors to this model (Supplemental Figure 5B and Supplemental Table 4.5), mirroring prior research that associated gut microbial BCAA synthesis genes with T2D in adults (34) and children with obesity (35). No factors were significantly associated with HOMA-IR score at baseline (data not shown).

### Donor-status independent metabolites and microbes were associated with weight gain and percent adipose in FMT recipient mice

Previous FMT experiments demonstrated that microbial content can confer phenotypic and metabolomic traits of the donor, at least temporarily, to germ-free mouse (19–21) and human (36) recipients. However, all prior studies that examined the role of the microbiota in obesity used fecal samples from adult donors. We hypothesized that adolescent microbiota, influenced by host growth and development, are optimized for peak energy harvest regardless of OB and HWC status, and that weight gain may not differ between mice receiving FMT from HWC or OB donors. To test this, we chose FMT samples of male and female donor adolescents from the HWC group, with %95^th^ BMI that ranged from the 20^th^ to the 80^th^ percentile, and from the OB group (see Supplemental Table 5 for FMT donor demographics). Male C57Bl/6 mice aged 5-7 weeks were weighed, gavaged with 150-200 μl of donor slurry (Figure 5A), housed under gnotobiotic conditions and fed a sterilized standard chow diet ad libitum, and then euthanized at 2 weeks post gavage. We weighed the ex-germ-free mice every 3-4 days post gavage (Figure 5B). While there was a non-significant trend towards more weight gained (Figure 5D) and heavier epididymal fat pads (Figure 5E) in mice that received donor slurry from adolescents with OB, we did not observe significant changes in weight gain or fat pad mass associated with donor OB vs HWC status or with sex or race of donor (data not shown). The strongest predictor of weight gain across all recipient mice was the starting weight of the recipient (R^2^=0.72, p<0.0001). Comparing metabolite profiles across all recipient mice, levels of serum amino acids Gly, Tyr, Asx, and Arg, as well as acylcarnitines C16:1, C10:3, C8:1C4-DC/Ci4-DC were negatively associated with weight gained (Figure 5C).

**Figure 5.**
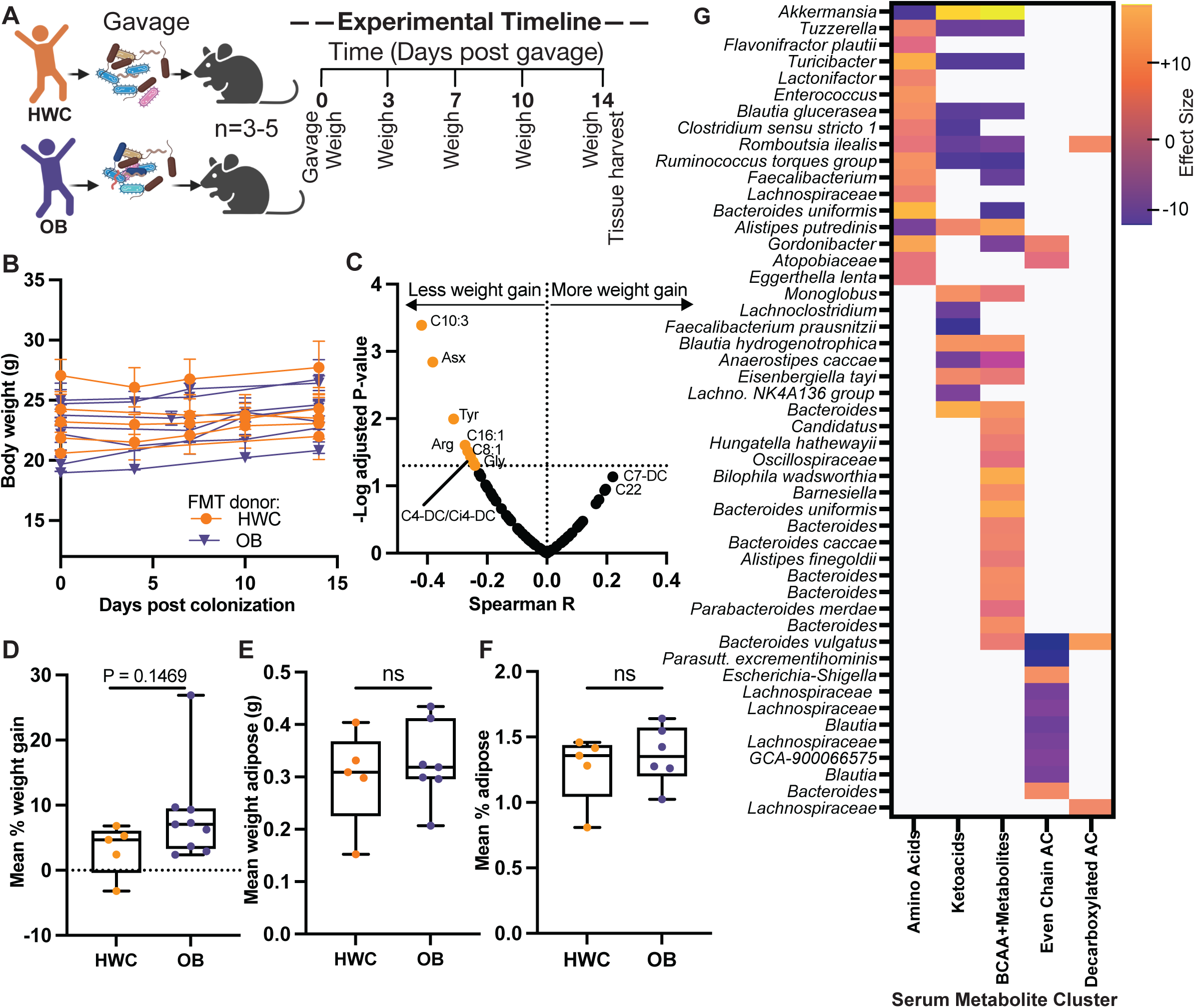
Fecal microbial transplants from adolescent POMMS study participants did not significantly affect weight gain in recipient mice but did influence serum metabolites. **(A)** Experimental design. Groups of 3-5 six-week-old male germ-free mice were weighed and gavaged with a slurry of baseline fecal material from one of 5 HWC or 9 OB POMMS participants. Mice were weighed every 3-4 days and sacrificed at 14 days post gavage. **(B)** Average weight and standard deviation of each group of mice over the 2 weeks post gavage period. **(C)** Mouse serum metabolites associated with percent weight gain over the FMT incubation period. Labeled dots above the dashed line represent metabolites significantly associated with less weight gain over time following linear regression analysis. **(D)** Mean percent weight gain of all mice grouped by donor (HWC v OB) at baseline. **(E)** Mean epidydimal fat pad weight grouped by donor at baseline. **(F)** Mean percent epidydimal fat pad weight grouped by donor at baseline. **(G)** Fecal microbial taxa were identified by 16S rRNA gene sequencing. ASV significantly associated (P < 0.05 after adjustment for multiple comparisons) with clustered mouse serum metabolites (Supplemental Figure S6) using ANCOM-BC (32) are shown on the heatmap. Clusters are described by the predominant metabolite species in each cluster. Each taxon in the heatmap represents a unique ASV identified to the lowest classification possible. BCAA, branched chain amino acids; AC, acylcarnitines. Diagram in part **(A)** was created with BioRender.com.

We were intrigued to see that microbiome samples from some individual adolescent donors were able to confer greater weight gain over others, raising the possibility that some transplanted human gut microbes might be associated with weight gain. Indeed, we did detect a microbial signature associated with weight gained, percent adipose (Table 2), and serum metabolite features. Fecal microbial alpha diversity was significantly higher in mice receiving donor microbiomes from HWC donors than OB donor recipients (Supplemental Figure 6C). We found Bray-Curtis dissimilarity to be significantly associated with percent weight gain and donor weight group (p-values <0.01) but not adipose percent change (p-value >0.1). We also saw significant engraftment of *Akkermansia*, *Ruminoccocus*, and *Bacteroides* in fecal samples 2 weeks post gavage when compared to the composition of the donor sample, and a relative decrease in members of the *Blautia* genus throughout engraftment (Supplemental Figure 6B). Only two taxa were significantly negatively associated with weight gain across all recipient mice – ASV 153 *Barnesiella* sp. (log2 fold change −0.80, padj. 5.2^-13^) and ASV 21 *Collinsinella aerofaciens* (log2 fold change −0.19, padj 0.02). We also found several microbes significantly associated with clusters of co-varying metabolites (Supplemental Figure 6C and Supplemental Table 5.2). Many of these microbes are significantly associated with metabolite cluster 1, comprised of mostly even medium- to long-chain acylcarnitines. Cluster 2 was defined by the BCAA valine, BCKA KIC, KIV, and KMV, along with BCAA metabolite 3-HIB (37). This metabolite cluster was both positively and negatively associated with several *Blautia* and *Bacteroides* ASVs (Figure 5G).

**Table 2.** Gut microbial taxa significantly associated with percent adipose tissue in mice at 2 weeks post FMT.

| Family | Genus | Species | ASV | Effect Size | P adj* |
| --- | --- | --- | --- | --- | --- |
| Lachnospiraceae | *Hungatella* | *hathewayii* | ASV_51 | 3.59 | 0.001 |
| Oscillospiraceae | *Colidextribacter* | NA | ASV_203 | -2.90 | 0.05 |
| Eggerthellaceae | *Gordonibacter* | NA | ASV_200 | -2.71 | 0.02 |
| Lachnospiraceae | NA | NA | ASV_119 | 15.78 | 0.02 |
| Akkermansiaceae | *Akkermansia* | NA | ASV_4 | 3.06 | 0.05 |

\*P values adjusted for multiple comparisons

### Serum levels of aromatic and branched chain amino acids are positively associated with BMI and IR over the 6 month observational study period while levels of even chain acylcarnitines are negatively correlated with change in BMI and IR

We next sought to determine whether metabolic shifts co-occur with improvements in health metrics during weight loss intervention. Using adjusted linear regression models, we examined whether a change in metabolites correlated with a positive or negative change in BMI, HOMA-IR, or CRP. We observed that, in general, changes in aromatic and branched chain amino acids were positively associated with change in BMI, HOMA-IR, and HbA1c over the 6 months observational study period, while changes in several even chain acylcarnitines and glycine were negatively associated with changes in BMI and HOMA-IR. While the majority of participants either maintained or reduced their %95^th^ BMI score, HOMA-IR increased for most participants over the observation period (Figure 6A-B and Supplemental Figure 8) which is a common phenomenon during adolescence (38–40). While amino acids were not associated with CRP, several acylcarnitine metabolites were positively associated with CRP. Changes in these acylcarnitines were negatively associated with changes in BMI or HOMA-IR (Figure 6DF).

**Figure 6.**
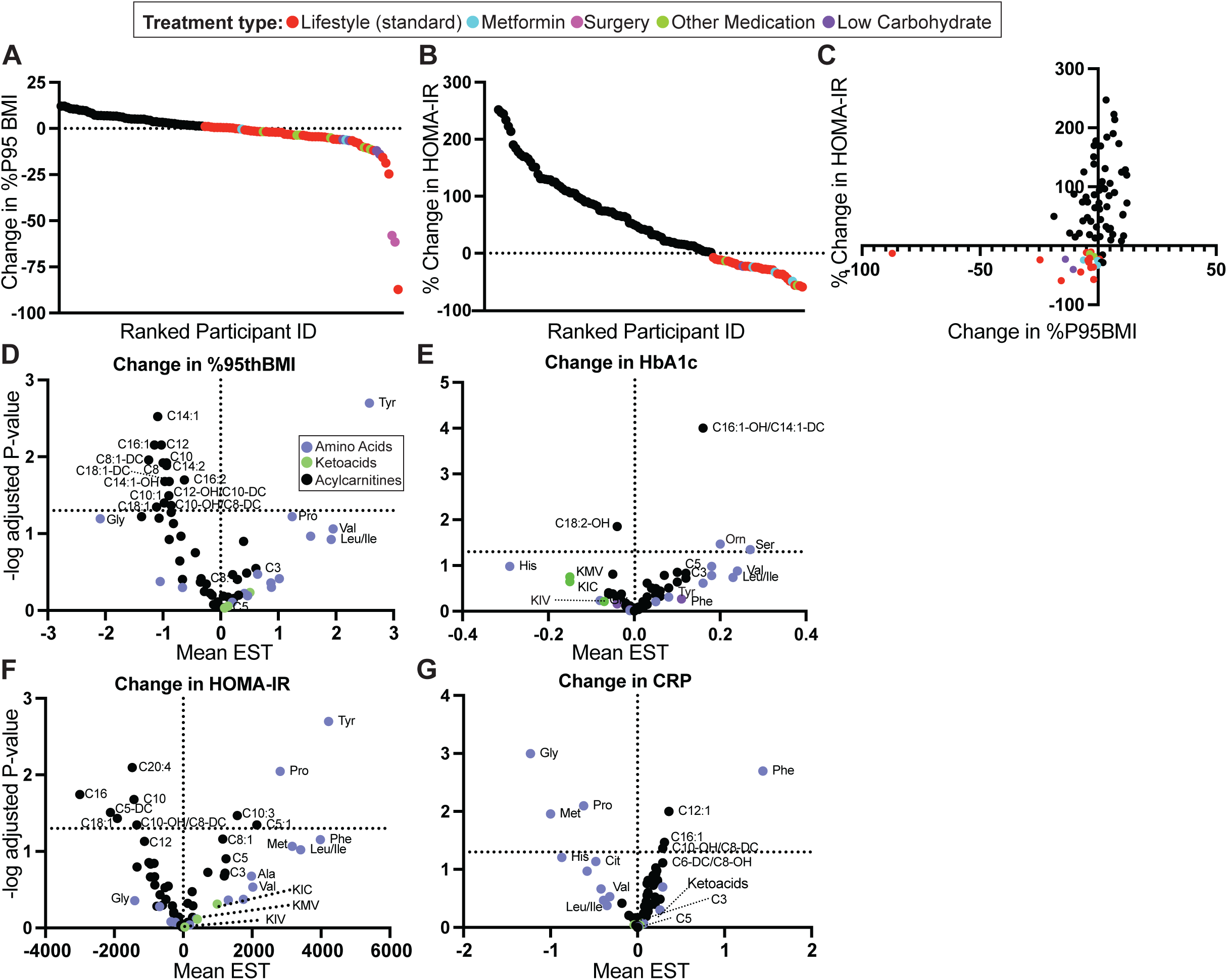
Serum aromatic amino acids increase with BMI and HOMA-IR scores over the 6 month observational study period, while several even chain acylcarnitines and glycine go up with clinical improvements and reduction in CRP. **(A)** Absolute change in %95th BMI in OB cohort participants over the observational study period. **(B)** Percent change in HOMA-IR over the observational study period. **(C)** Graph of change in %95th BMI and percent change in HOMA-IR over the observational study period. **(D-F)** Estimated coefficients for metabolite levels associated with change in %95th BMI **(D),** Percent change in HOMA-IR **(E)**, or change in CRP **(F)** after adjusting for age, race, sex, and baseline measurement. For **(A-C)**, all participants with net reduction in the indicated measure are labeled by observational trial treatment type. For **(D-F)**, acylcarnitines are labeled in black, amino acids are labeled in purple, and ketoacids are labeled in green.

### Microbiome taxa and pathways at baseline predict change in health measures over the observational period, while distinct sets of taxa and features are associated with change in CRP

We analyzed the metagenomic data to determine if there were any microbial taxa or pathways at baseline that were significantly associated with change in BMI, HOMA-IR, CRP, or HbA1c. While there were no significant associations with change in HOMA-IR, we found that relative abundance of several microbial taxa at baseline were predictive of changes in BMI, HbA1C, or CRP levels, with some taxa shared between changes in 2 measures (Figure 7B). For example, *Bacteroides thetaiotaomicron*, *finegoldii*, and *uniformis* as well as *Parabacteroides distasonis* and *merdae* were all associated with change in CRP and %95^th^ BMI, but often in opposite directions. In addition, *Anaerostipes hadrus*, *Blautia producta*, and *Escherichia* virus Lambda_4A7 were all commonly associated with a change in HbA1c (Figure S7C) and CRP (Figure S7D). Notably, a high number of taxa (93) were significantly associated with changes in CRP suggesting that the subset of microbes that potentially predict changes in this inflammatory marker are more abundant when compared to those associated with strictly with BMI or other metabolic health indicators.

**Figure 7.**
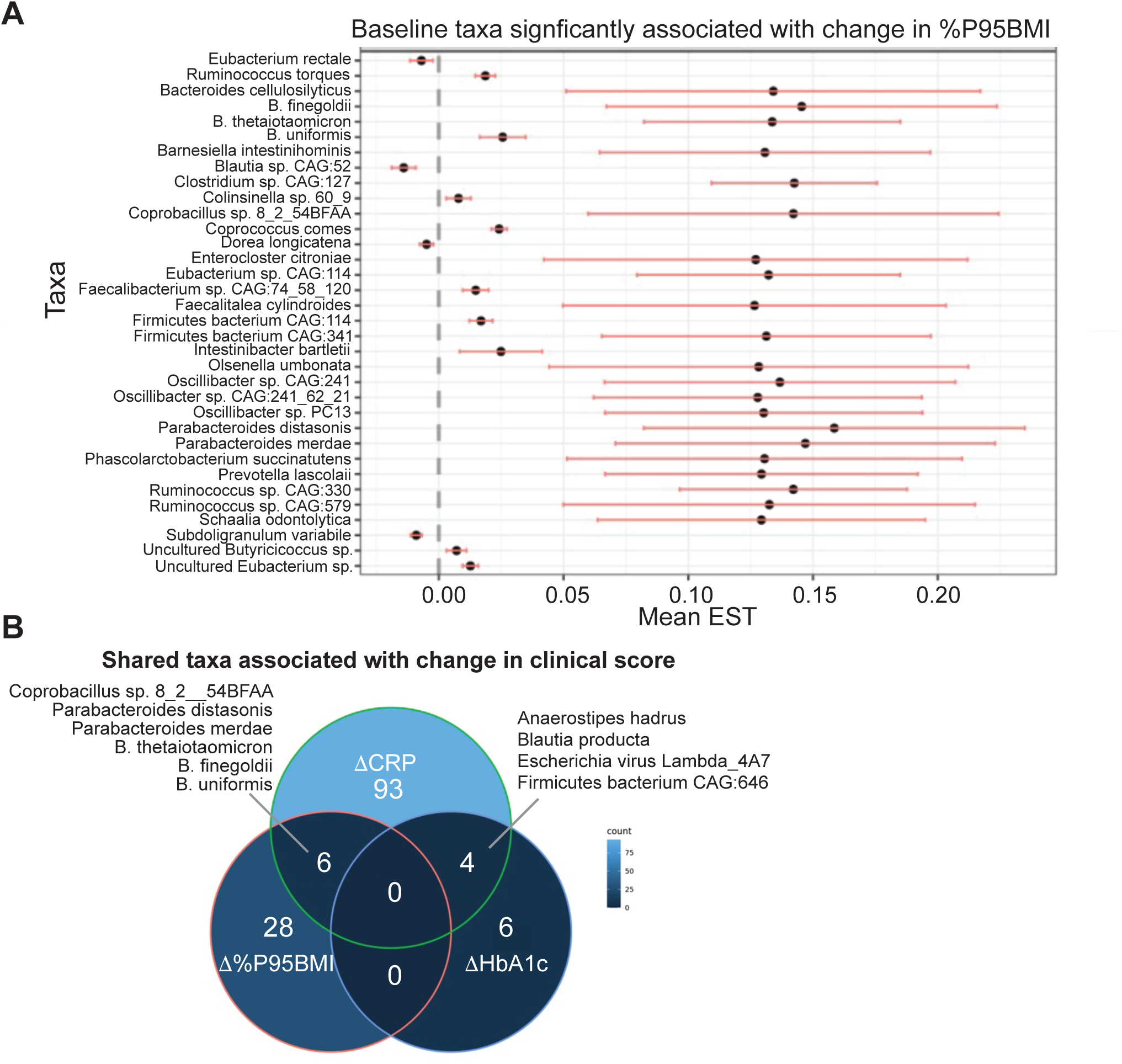
Microbial taxa associated with longitudinal outcomes. **(A)** Differential abundance of microbial taxa at baseline associated with change in %95th BMI over the 6-month observational period. **(B)** Venn diagram of microbial taxa shared between changes in clinical outcomes over the study period. See Statistical Analysis methods for details.

**Figure 8.**
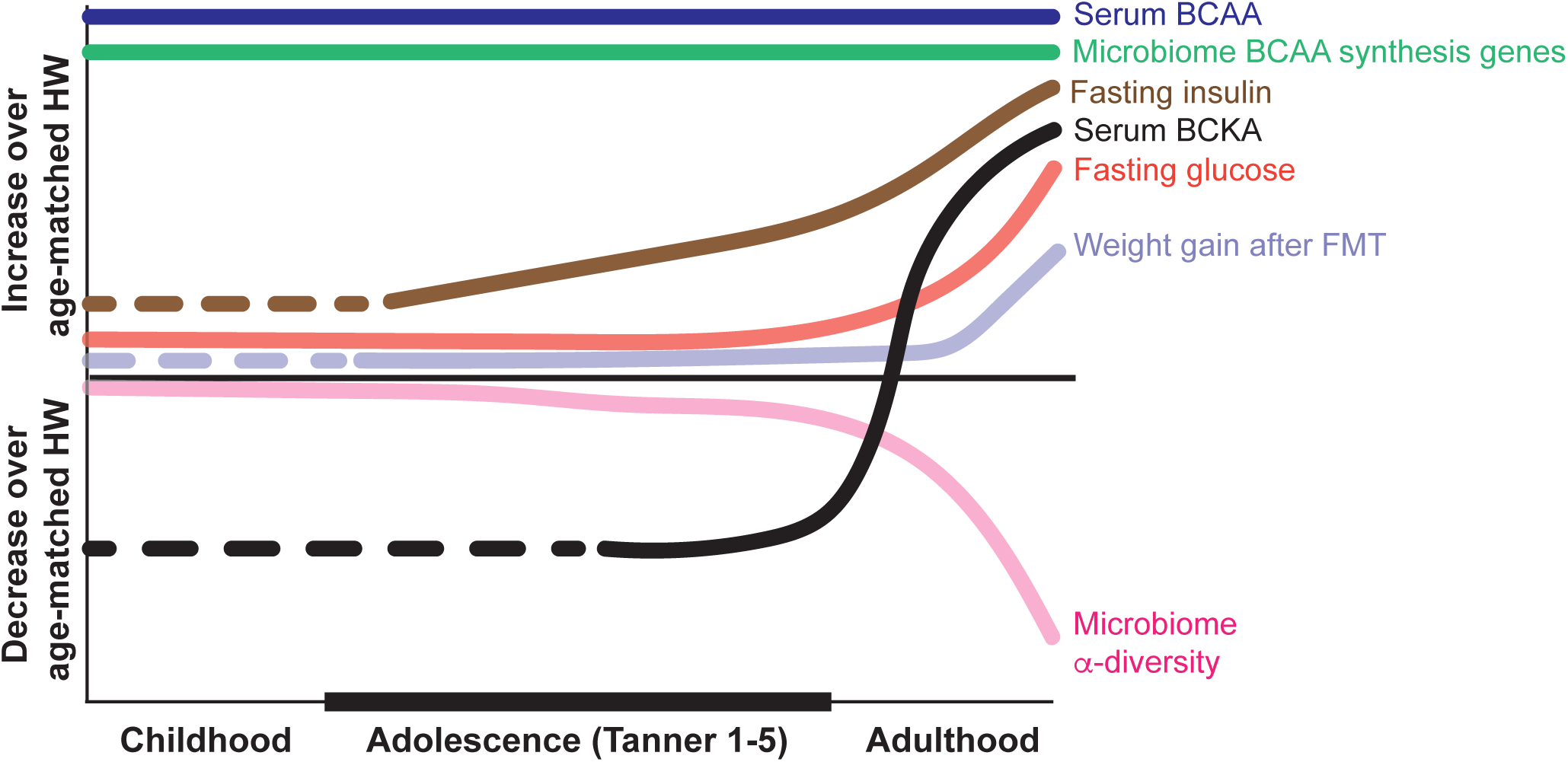
Obesity in adolescence is characterized by unique and potentially transitional metabolomic and microbiome features. Lines indicate the direction of microbiome and metabolome features in individuals with obesity relative to age matched healthy weight (HW) individuals. Several of these features display changing relationships with OB status across the lifespan. Dashed lines indicate age ranges where there is currently missing data for that feature. We found that while high BMI is associated with high levels of serum BCAA, BCKA differences between OB and HW adolescents are sex dependent and pronounced in later stages of puberty. Further, serum measures of insulin resistance increased during adolescence for nearly all participants during our observational study, regardless of whether the participant lost weight, but there was no difference in fasting glucose between OB and HWC at baseline. We saw no significant difference in the ability of adolescent microbiome to transmit more weight gain to germ free mice, while evidence suggests that microbiome transplant from adult donors with OB leads to more weight gain in recipients. Evidence for each of these associations is cited in the text.

## DISCUSSION

Based on studies of adult obesity indicating that the gut microbiome plays a substantial role in driving obesity-associated phenotypes, similar microbiome and metabolic coupling has been presumed in adolescent obesity. We studied a cohort of adolescents with and without obesity to compare their metabolic and microbiome states with the hypothesis that the obesity-associated adolescent metabolome and microbiome differ from that reported in adults with obesity. We further postulated that the adolescent metabolic rearrangement associated with obesity would not be associated with gut dysbiosis to the same extent as observed in adults, suggesting a decoupling of metabolic shaping in adolescent obesity and the gut microbiome.

### BCAA and BCKA have an inversely correlated signature in obesity that is unique to adolescents

In our study of adolescents with obesity, we found that serum levels of the BCAA and their direct metabolites, the BCKA, significantly differed between the OB and HWC cohorts, but the BCKA skewed higher in the HWC group. At the same time, BCAA were higher among adolescents with obesity. Circulating levels of BCAA have been associated with obesity, IR, and T2D in adults since the late 1960s (1, 2, 41). At baseline, BCAA and aromatic amino levels are prognostic for the development of T2D in adults in long-term studies (3, 42), and this same metabolite cluster also predicts improvements in HOMA-IR in response to weight loss (43). While circulating amino acids are often measured in metabolite studies of adults, it is less common that those data sets also include BCKA measurements. When measured, BCKA levels tend to mirror BCAA levels in adults with obesity, with both being higher than in HWC (7) and both dropping significantly after effective treatments for obesity and T2D such as an exercise-based intervention (6) or bariatric surgery (44, 45). This may be related to obesity-driven changes in expression and activity of the kinase (branched-chain ketoacid dehydrogenase kinase, BCKDK) and phosphatase (protein phosphatase Mg2+/Mn2+ dependent 1 kinase, PPM1K) enzymes that control phosphorylation state and activity of branched-chain ketoacid dehydrogenase (BCKDH), the rate-limiting enzyme of BCAA catabolism (46, 47). Similarly, studies in adult rodent models, including *ob*/*ob* mice and Zucker-obese rats, show combined higher levels of BCAA and BCKA in the obese state (7, 47). Our finding of anticorrelated BCAA and BCKA levels in adolescents with obesity indicates a metabolic phenotype during these critical early stages that is distinct from that found in adults with obesity. We speculate this could be due to flexible regulation of PPM1K/BCKDK in adolescents allowing for BCAA catabolism even with higher circulating BCAA, or patterns of organ-specific BCAA metabolism in adolescents that are distinct from what has been observed in adults. In agreement with our results, a recent study in adolescents suggested that high BCAA and low Gly should be included as part of a metabolic signature that could predict β-cell failure (48).

We found that the intersection of sex and obesity status plays a significant role in BCAA and BCKA levels. Our data suggest that these effects do not emerge until puberty onset, as we found that there were few sex-based differences in BCAA-BCKA metabolites in kids aged 5-9 years from the AHA Hearts and Parks Study (30). Our observation of sexual dimorphism in circulating levels of BCAA and their metabolism in adolescents is in accord with previous studies. Kobayashi and colleagues demonstrated that female sex hormones affected the diurnal regulation of the enzyme complex responsible for BCAA metabolism in rat liver, whereas gonadectomy of adult male rats did not affect circadian BCAA metabolism (8). Human data examining the serum metabolomic differences of 1756 adult participants (903 females and 853 males) from the population-based Cooperative Health Research in the Region of Augsburg Germany (KORA) F4 study suggested that BCAA metabolites were significantly higher in males and were significantly associated with male sex when taken as a group (49). In another study, exercising men had higher rates of leucine oxidation than women (50). Focusing on children and teens, Newbern and colleagues (51) published that in a cohort of 82 teens with obesity, BCAA levels and byproducts of BCAA catabolism are higher in teen boys with obesity than in girls of comparable BMI. A subsequent study by the same group found that plasma levels of KMV, the ketoacid of isoleucine, went down in males but not in females when measured at the 6-month follow-up visit (16). The mechanisms underlying these emerging sex- and age-associated differences in BCAA metabolism remain unknown, but they could represent useful diagnostic and prognostic markers of obesity-associated pathologic progression.

### Lower serum glycine is a common signature of obesity in adolescents and adults

We also found that serum glycine levels were significantly lower in adolescents with obesity when compared to the HWC group. This observation has been documented in adult obesity (52–54), and glycine levels increase after weight loss and improvement with insulin sensitivity (6, 44, 45, 55, 56). A recent study showed that elevations of BCAA in obesity activate glycine consumption to replenish pyruvate needed for the alanine transaminase reaction and nitrogen unloading in skeletal muscle (57) (Figure 2A). Treatment with a specific BCAT inhibitor that interferes with BCAA transamination increases muscle and plasma glycine levels in Zucker fatty rats (10). Our data suggest that the reciprocal relationship between BCAA catabolism and glycine levels exists in adolescents despite their divergent BCKA signatures.

Acylcarnitine data from adult humans suggests that serum or plasma levels of C3-C5 species, which are derived from the metabolism of BCAA and other amino acids (58), often cluster in principal component metabolite factors along with the BCAA and aromatic amino acids, with higher levels in obesity, insulin resistance, and T2D (2, 43, 59). Low-grade chronic inflammation is a hallmark of T2D (60, 61) and obesity (62), and CRP levels drop with weight loss (63, 64). Herein, we identified an acylcarnitine signature that included associated with CRP levels at baseline (Figure 1) and with change in CRP during the observational period (Figure 6) that was distinct from the acylcarnitine species associated with both baseline and during-study changes in BMI and HOMA-IR. Fewer data sets incorporate metabolites when comparing HWC and OB in children and adolescents. One of the most comprehensive reports describes a cohort of 803 Hispanic children in the Viva la Familia study, ranging in age from 4-19y and split between HWC and OB at a single timepoint (15). They examined 304 named serum metabolites and found that their OB subjects had increased BCAA and acylcarnitine metabolites and increased inflammation markers. The Viva la Familia study did not include BCKA measurements, nor did it include longitudinal sampling or interventions. The data sets reported here therefore provide new insights into the relationships between BCAA, BCKA, and acylcarnitine species in the context of pediatric obesity and intervention outcomes.

### Functional microbiome signatures are associated with obesity and metabolic signatures

Fecal microbiome transplants from adult lean donors and donors with obesity into germ-free mice demonstrate that the microbiome can contribute to weight gain and adiposity and affect the serum metabolomic profiles of the recipients (19, 21, 65). While we did not observe significant differences in alpha and beta diversity between our adolescent case and control groups, the functional suite of genes represented in the microbiome associated with obesity in adolescence may still play a role in the serum availability of some obesity-related metabolic biomarkers. We found significant associations between BMI status and microbial genetic ability to synthesize several amino acids including the BCAA and tryptophan (Supplemental Figure 5B). Research in pigs suggests that approximately 30% of dietary glycine is consumed by the small intestinal microbiota (66, 67), while other work in both conventionally-raised and germ-free mice and rats suggests that the gut microbiome contributes on the order of grams per day of essential amino acids to the nutritional pool (65). In the future, longitudinal studies in pre-clinical models of diet-induced obesity could be used to clarify the role of age and sex in BCAA and acylcarnitine metabolism, to investigate mechanisms underlying the transition from high BCAA - low BCKA in youth to high BCAA - high BCKA in adult obesity, and the contributions of the microbiome to those metabolic shifts.

### Metabolites and microbiome associations in children and adolescents – microbiome across HWC and OB adolescents is less differential than in adult cohorts

Our results provide important insights into the composition and function of the gut microbiome of pediatric obesity. Previous studies suggest that the gut microbiome undergoes changes between adolescence and adulthood in healthy individuals (68). Adults with obesity and healthy weight display substantial differences in gut microbiome diversity and composition (19, 69, 70), and those differences are distinct from adolescents with obesity or healthy weight (71). Our comparisons of adolescents with obesity and healthy weight detected no differences in diversity and limited differences in composition and functional potential. Previous studies examining the role of the microbiome in the development of obesity in germ-free animal models demonstrated that the microbiome from adult donors with OB contributes to more weight gain, higher adiposity, and higher circulating BCAA compared to a microbiome transplant from a lean adult donor (19, 72). However, there was no specific association between weight gain and donor BMI in our FMT studies here from adolescent donors. Together, these results raise the possibility that the gut microbiomes harbored by adolescents with obesity do not yet have sufficient compositional or functional changes to causally affect host metabolism, whereas microbiome changes and causal consequences increase progressively as individuals with obesity progress into adulthood. In the future, it will be important to determine if the ability of the gut microbiome to contribute to obesity phenotypes is due to microbiome functional differences (69) or differences in host growth and physiology during that critical stage. The resulting information could inform efforts to target the microbiome in treatment of obesity.

Recently, several medications were FDA-approved for the treatment of obesity in adolescents aged 12 and older (24). While these agents are remarkably effective in reducing body mass index (BMI), questions remain about the necessary duration of treatment. Some studies have suggested that a normalization of BMI prior to adulthood, in effect, “resets” the metabolic pathways and may offer hope for these teens to avoid the need for lifelong medication. In the future, it will be interesting to determine how these newly approved medications affect the distinct metabolic and microbial phenotypes described here.

One limitation of this study is that we did not have longitudinal sampling of any HWC adolescent to examine any changes in insulin resistance associated with typical adolescence to compare to adolescents with OB in our cohort (38–40). Another limitation is that our linear regression data associations between metabolites and clinical values were only adjusted for demographics and not multiple comparisons, so while relationships seem strong, the reported P values are nominal to generate hypotheses that could be further evaluated in murine functional models.

In conclusion, we found that adolescents with OB have unique metabolomic adaptations when compared to their HWC counterparts and adults with OB. We also found that while both OB and HWC teens had similar measures of gut microbiome diversity, there were distinct microbial taxa and functional differences that defined OB v HWC status. We also found that the suite of metabolites and gut microbial taxa associated with inflammatory status as measured by CRP level had little overlap with those associated with insulin resistance scores and BMI. Longitudinal analysis revealed metabolic and microbial features that correlate with or predict changes in health measures over the observational period. Data derived from this cohort will help to develop an improved understanding of the physiological pathways of adolescent obesity and may provide insights that will inform future clinical decision-making.

## METHODS

### Sex as a biological variable

Both male and female participants were included in this study. All relevant analyses were adjusted for sex, age, and race.

### Study approval

Recruitment of participants and handling of clinical samples was approved as part of the Pediatric Obesity Microbiome and Metabolism Study (POMMS; Clinical trial identifier # NCT03139877) and described in detail in (23). Written informed consent was received prior to participation and prior to inclusion of serum and fecal samples for further study in the POMMS biobank. Data, serum, and stool samples from the study participants have been banked and are available as a repository (Clinical trial identifier #NCT02959034) for further research as described at https://sites.duke.edu/pomms. All animal experiments were performed either at the Duke University Gnotobiotic Core or in the National Gnotobiotic Rodent Research Center at the Center for Gastrointestinal Biology and Disease at the University of North Carolina Chapel Hill in AAALAC-accredited facilities and according to IACUC-approved protocols.

### Serum metabolomics

Serum metabolomic samples from the Hearts and Parks study (30) were randomized and analyzed with the POMMS cohort samples to reduce drift from batch effects. All serum metabolomics assays were performed at the Duke Molecular Physiology Institute. Serum samples were randomized and analyzed for the conventional metabolites nonesterified fatty acids (NEFA), glucose, insulin, glycerol, blood urea nitrogen (BUN), 3-hydroxybutyrate (3-HB), total ketones, triglycerides, and uric acid. The samples were run in seven daily batches of approximately 76 sera per batch. Samples were run on a Beckman autoanalyzer from individual tubes, with pooled serum controls inserted at the beginning, middle, and end of the sequence. Reagents for measuring for total ketones, 3-HB, and NEFA were from Fujifilm (Osaka, Japan). Samples were additionally assayed for insulin using kits from Meso Scale Discovery (Rockville, MD). All samples were refrozen afterwards for later insulin immunoassays on 96-well plates. Corresponding to each batch of metabolites were two insulin plates (e.g. 1-2 for Batch 1, 3-4 for Batch 2, and so on) on which the 76 samples were assayed, 38 per plate. Controls were run twice on each insulin plate, placed at the beginning and end. Insulin was measured by immunoassay using reagents and electrochemiluminescent imager from Meso Scale Discovery (Rockville, MD). Glucose was measured on a Beckman DxC 600 clinical analyzer (Brea, CA). HOMA-IR was calculated as (fasting insulin in units/liter x fasting glucose in mg/deciliter)/405 (73).

Serum samples were analyzed for amino acids and acylcarnitines in a targeted assay using labeled standards as described (23). BCKA were analyzed by LC-MS/MS as described (10). Briefly, 20 µl of plasma containing isotopically labeled internal standards KIC-d3, KIV-5C13 (Cambridge Isotope Laboratories), and KMV-d8 (Toronto Research Chemicals) was precipitated with 150 µl of 3M PCA. 200 µl of 25 M *o*-phenylenediamine (OPD) in 3M HCl was added to the supernatants, and the samples were incubated at 80°C for 20 minutes. Keto acids were extracted with ethyl acetate as previously described (74). The extracts were dried under nitrogen, reconstituted in 200 mM ammonium acetate, and analyzed on a Waters Xevo TQ-S triple quadrupole mass spectrometer coupled to a Waters Acquity UPLC system. The analytical column (Waters Acquity UPLC BEH C18 Column, 1.7 μm, 2.1 × 50 mm) was used at 30°C. 10 μl of the sample was injected onto the column and eluted at a flow rate of 0.4 ml/min. The gradient consisted of 45% eluent A (5 mM ammonium acetate in water) and 55% eluent B (methanol) for 2 min., followed by a linear gradient to 95% B from 2 to 2.5 min., held at 95% B for 0.7 min., returned to 45% A, and then the column was re-equilibrated at initial conditions for 1 minute. The total run time was 4.7 min. Mass transitions of *m/z* 203 → 161 (KIC), 206 → 161 (KIC-d3), 189 → 174 (KIV), 194 → 178 (KIV-5C13), 203 → 174 (KMV), and 211 → 177 (KMV-d8) were monitored in a positive ion electrospray ionization mode.

### Fecal DNA preparation and sequencing

Fecal DNA was collected and aliquoted as described (23). DNA was recovered from approximately 200mg of fecal material using the DNeasy Power Soil Pro kit (Qiagen). Bacterial community composition in isolated DNA samples was characterized by amplification of the V4 variable region of the 16S rRNA gene by polymerase chain reaction using the forward primer 515 and reverse primer 806 following the Earth Microbiome Project protocol (http://www.earthmicrobiome.org/). These primers (515F and 806R) carry unique barcodes that allow for multiplexed sequencing. Concentration of the PCR products was measured using a Qubit dsDNA HS assay kit (ThermoFisher, Q32854) and a Promega GloMax plate reader. Equimolar 16S rRNA gene PCR products for up to 384 samples were pooled prior to sequencing. Sequencing was performed by the Duke Sequencing and Genomic Technologies (SGT) Shared Resource on an Illumina NovaSeq instrument configured for 250 base-pair paired-end sequencing runs on a S Prime lane. Fecal DNA extraction, 16S rRNA library preparation, and sequencing, was performed at the Duke Microbiome Core laboratories. Taxa were identified using DADA2 (75) and resulting data was analyzed as described in the Statistical Analysis section.

Fecal DNA from a subset of 396 samples was submitted for metagenomic sequencing at the Novogene Corporation, Inc., using the following protocol. Genomic DNA was randomly fragmented by sonication, then DNA fragments were end polished, A-tailed, and ligated with the full-length adapters of Illumina sequencing, and followed by further PCR amplification with P5 and indexed P7 oligos. The PCR products as the final construction of the libraries were purified with AMPure XP system. Then libraries were checked for size distribution by Agilent 2100 Bioanalyzer (Agilent Technologies, CA, USA), and quantified by real-time PCR to meet the criteria of 3 nM. The qualified libraries were fed into Illumina NovaSeq sequencers after pooling according to effective concentration and expected data volume. The raw sequenced reads were then filtered to remove reads that contained adapters, reads containing greater than 10% unidentified bases, and reads with a quality score <5, or over 50% of the total bases. This resulted in a total output of 2871.0 Gb of raw data.

### Fecal microbiome transplant experiments

Gnotobiotic experiments were conducted at the Duke University Gnotobiotic Core Facility or the National Gnotobiotic Rodent Resource Center in the Center for Gastrointestinal Biology and Disease at the University of North Carolina at Chapel Hill. Human fecal samples were prepared for animal gavage as described (76). Briefly, upon receipt, samples were initially frozen at −80°C and then thawed, opened in a flexible film anaerobic chamber (Coy Laboratory Products, Grass Lake, Michigan), homogenized with a sterile plastic loop, and distributed into 2 ml cryovials in ∼200 mg aliquots without preservative. Samples were then stored at −80°C. Immediately before fecal transplant, samples were thawed, resuspended into 10% weight per volume sterile PBS +0.05% cysteine, vortexed repeatedly, allowed to settle at room temp for 5-10 min, and supernatant was then removed for gavage. Three, four, or five male germ-free C57BL/6 mice between 5-7 weeks of age received between 150 and 200 μl of gavage slurry for each human sample administered. Mice were then housed with ALPHA-Dri (Shepherd Specialty Papers) bedding for two weeks in sterile filter top cages and fed autoclaved Teklad 2020SX diet (Envigo) and water ad libitum. Mice were weighed at gavage and every 3-4 days following fecal transplant. At 2 weeks, mice were euthanized by isoflurane overdose and immediately exsanguinated by cardiac puncture. Fecal samples and tissues were removed and immediately snap frozen in liquid nitrogen, with between 5- and 7-min elapsing between euthanasia and complete tissue removal. Serum and tissue targeted metabolites were measured as described above and as previously published (23).

### Statistical Analysis

Clinical lab measures and targeted metabolites were log-transformed to ensure normality. Any metabolite with missing values (i.e. below the lower limits of quantification) in >25% of samples was not further analyzed. For the remaining metabolites, values below limit of quantification were imputed at half the minimum observed result (77). Logistic regression models were used to test association between each clinical lab measure with OB vs. HWC groups, controlling for age, sex, and race/ethnicity. To evaluate the metabolic status of study participants, serum levels of 65 targeted metabolites were investigated for associations with baseline %95^th^ BMI, HOMA-IR score, HbA1c, and CRP using linear regression models adjusting for age, sex, and race/ethnicity. Next, we assessed whether the association between case-control status and each metabolite varied by age, sex, or race using linear regression model with the interaction between status and age, sex, or race in separate models. If the overall interaction was statistically significant (nominal p<0.05), pairwise comparisons were reported. Lastly, the association between change in each clinical outcome (%95^th^ BMI, HOMA-IR score, HbA1C, and CRP) and change in each metabolite between baseline and 6-months were assessed using linear regression models adjusting for age, sex, race/ethnicity, and baseline clinical outcome. All analyses were conducted in R 4.1 (R Core Team, 2022). Significance values were set at p<0.05 throughout. Microbiome data was analyzed as follows. Spearman correlation was used to assess the association of alpha diversity metrics (Observed species, Shannon index, Faith phylogenetic diversity) with mouse or human physiologic measures, and Wilcoxon rank sum test was used to determine significant differences between diversity metrics. Principal coordinate analysis with Bray-Curtis dissimilarity was used to determine differences in overall microbial composition between gavage and fecal samples as well as a clear clustering of fecal samples by donor and compositional differences were determined by PERMANOVA (R package vegan). Hierarchical clustering (R package hclust, default setting) was used to cluster metabolites as well as species in the metagenomic data set.

Taxa were assembled, annotated, and functionally characterized using Kaiju (78) and HUMAnN (79) pipelines. Differential abundance analysis of ASVs with relative abundance above 0.1% in at least 20% of the samples (R package ANCOMBC) was performed for donor weight group, percent weight gain, and adipose percentage change, respectively, as well as for metabolite factors jointly. The Benjamini-Hochberg method was applied to control the false discovery rate (FDR) at 0.05 for multiple comparisons of ASVs. Random forests machine learning was (80) used to model associations between fecal metagenomic taxa or functional genes and clinical measures.

## Supporting information

Supplemental tables with raw data

## Data availability

Raw unprocessed and log-transformed data can be found in Supplemental Data attached to this article. All raw sequencing data can be accessed via the NIH Sequence Read Archive at Accession number (**number forthcoming).** Any other data and R code will be shared upon request.

## ACKNOWLEDGEMENTS

The authors are grateful to Kristin Cleveland, Jamison Cameron, Brittany McAdams, and staff in the Duke Gnotobiotic Core in the Division of Laboratory Animal Resources in the Duke University School of Medicine, and Jeremy Herzog, Josh Marc Frost, and R. Balfour Sartor in the National Gnotobiotic Rodent Resource Center in the Center for Gastrointestinal Biology and Disease at the University of North Carolina at Chapel Hill which is supported by P30-DK034987. We also thank staff at the Duke Microbiome Core laboratories for their assistance in preparing samples for high throughput sequencing.

