## Supplements figures and legends for "Branched chain amino acid metabolism and microbiome in adolescents with obesity during weight loss therapy"

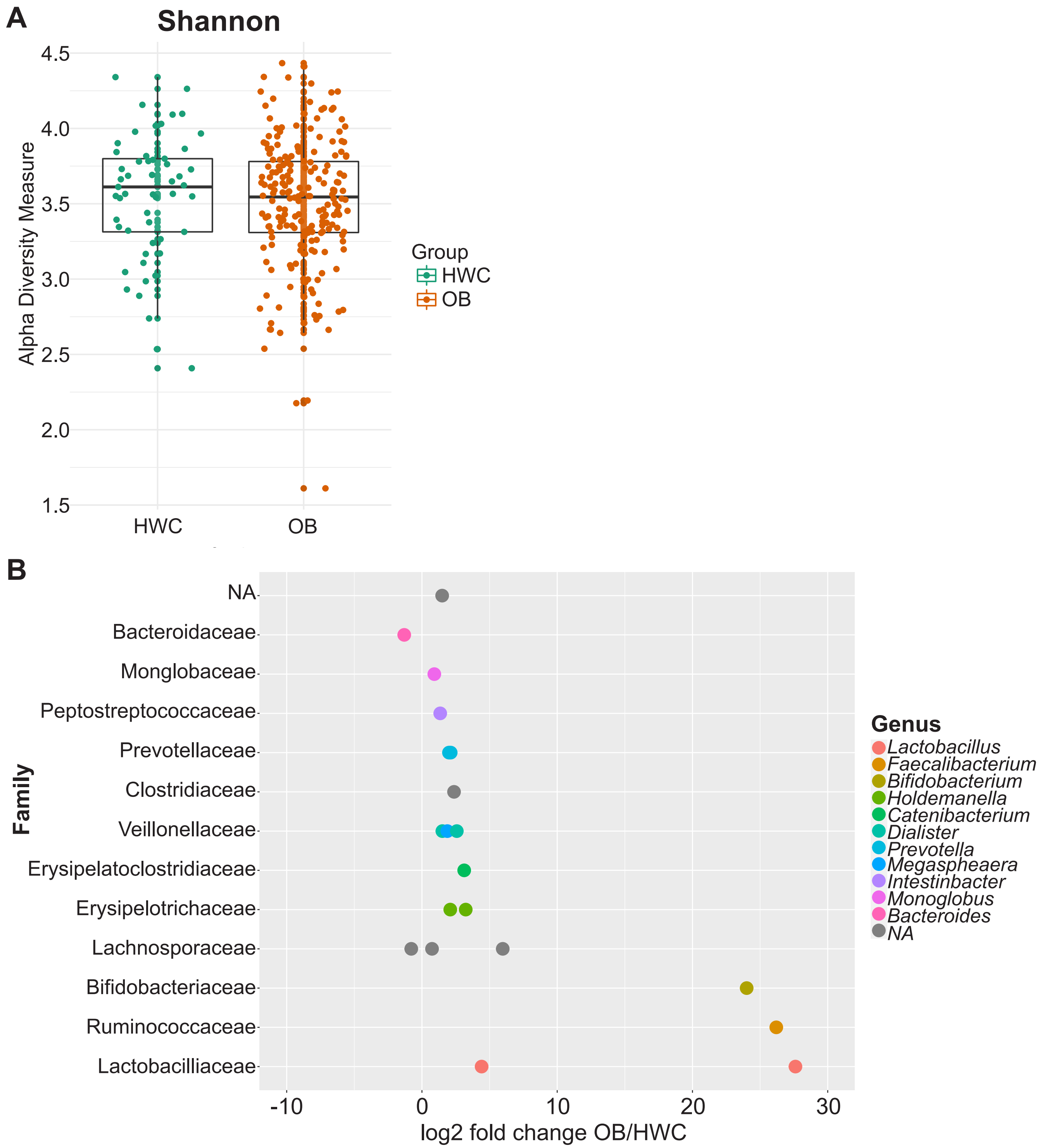


**Supplemental Figure 1. Gut microbiome diversity was similar between HWC and OB cohorts at baseline, but the abundance of several genera were significantly different between groups**. **(A)** Alpha diversity metric (Shannon index) of gut microbial taxa as measured by 16S rRNA gene high throughput sequencing. HWC, healthy weight control; OB, adolescents with obesity. **(B)** Deseq2 measure of differential abundance at the family and genus level, when available, of microbial taxa comparing OB and HWC groups.

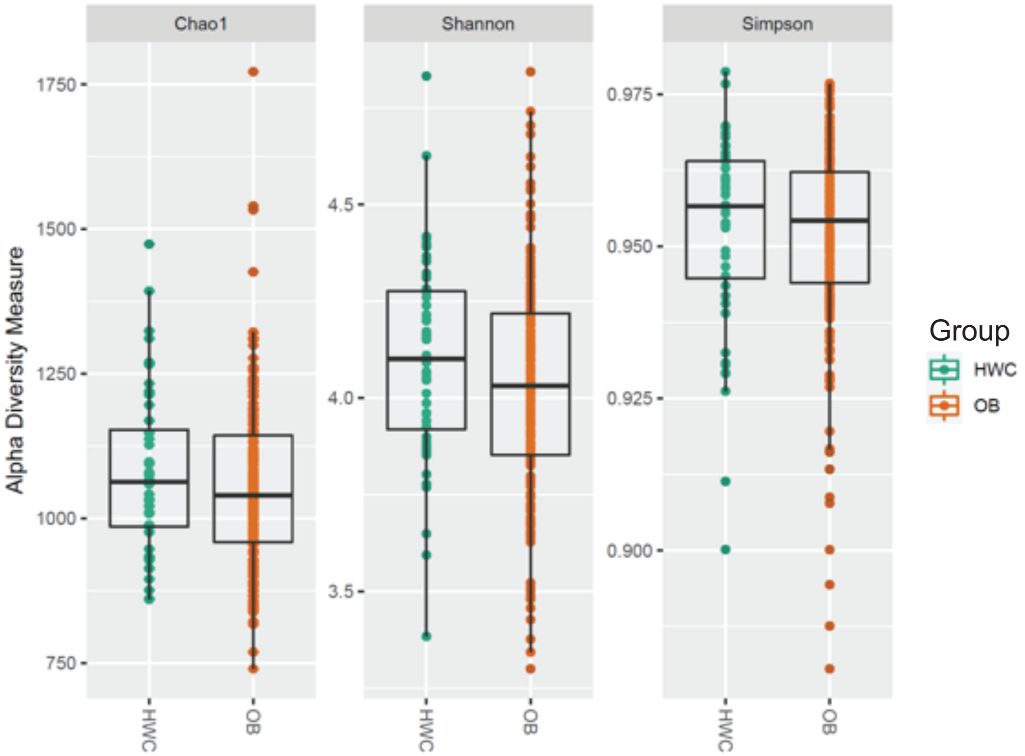


| | p-value | Kruskal-Wallis chi-squared |
|---|---|---|
| *Chao1* | 0.279628364997128 | 1.16889519261103 |
| *se.chao1* | NaN | NaN |
| *Shannon* | 0.118436952574392 | 2.43786996603581 |
| *Simpson* | 0.407967447520968 | 0.684716157205344 |

**Supplemental Figure 2. Shotgun DNA sequencing on a subset of fecal samples resulted in similar alpha diversity measures between OB and HWC cohorts at baseline**. A table of statistical analysis result values and p-values is also shown.

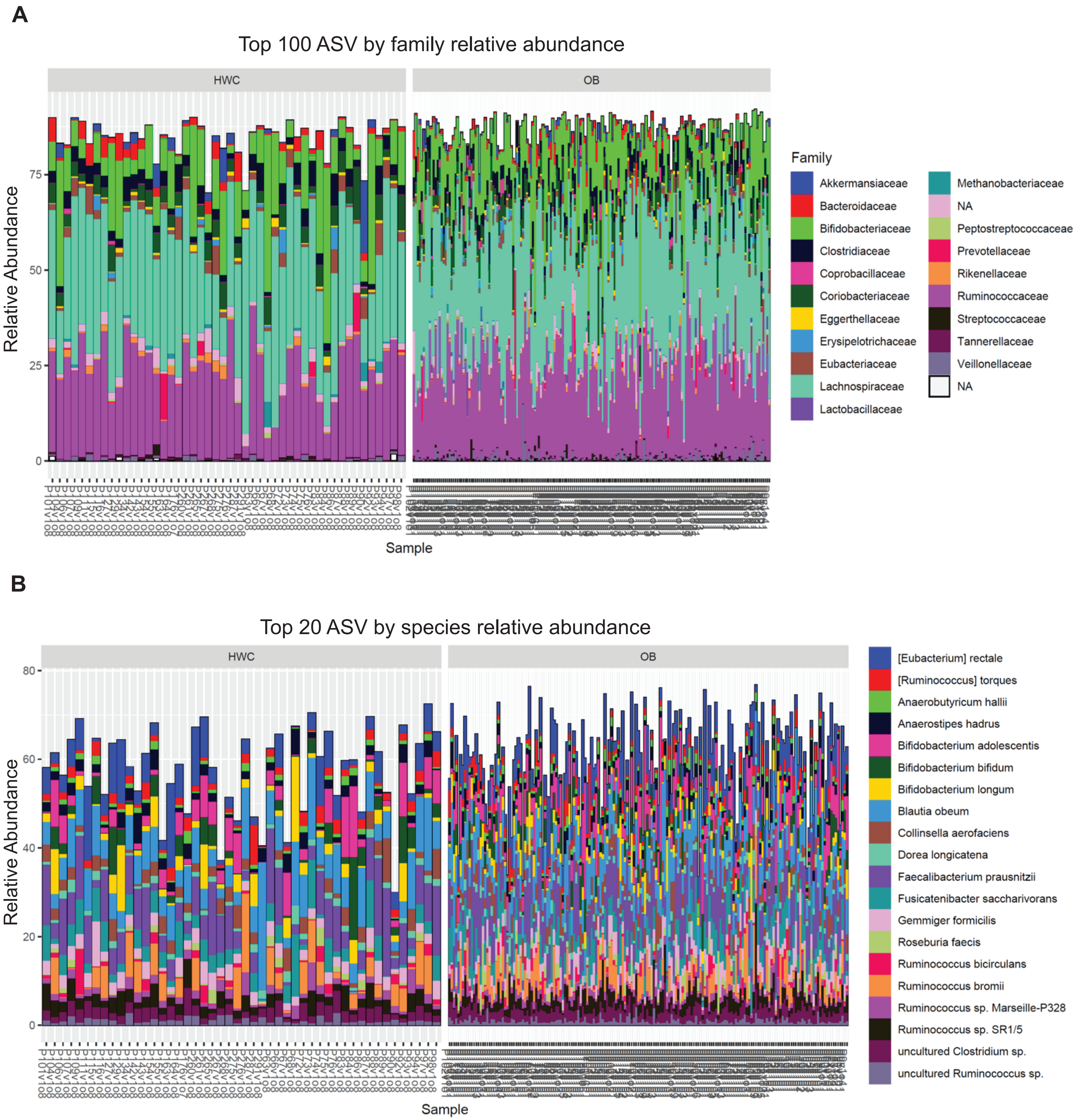


**Supplemental Figure 3. Shotgun DNA sequencing demonstrated similar species and family community structure between OB and HWC cohorts at baseline**. The top 100 most abundant families **(A)** and the top 20 most abundant species **(B)** from fecal metagenomic sequencing results are shown.

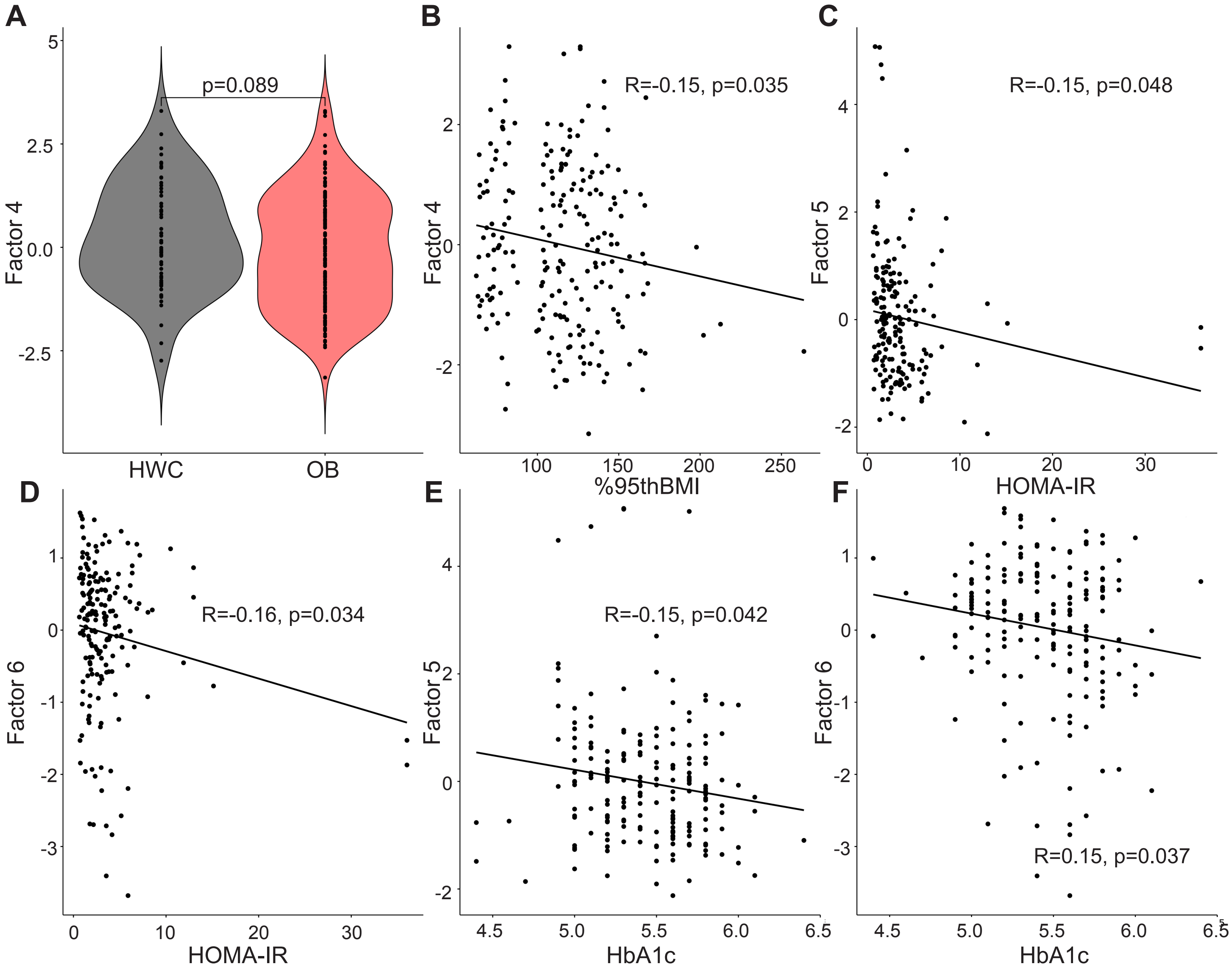


**Supplemental Figure 4. Factor analysis suggests clusters of co-varying microbial taxa and functional gene content associated with clinical measures of health.** Microbial taxa and gene content as defined by shotgun sequencing were clustered based on co-variance using hierarchical clustering. **(A)** Overall cluster member abundance from Factor 4 between HWC and OB cohort. Clusters with significant associations by linear regression with **(B)** %95thBMI, **(C-D)** HOMA-IR, or **(E-F)** HbA1c are shown. Taxa and functional components that making up each of the microbial factors are identified in "Supplemental Table 2 – Factor Weights."

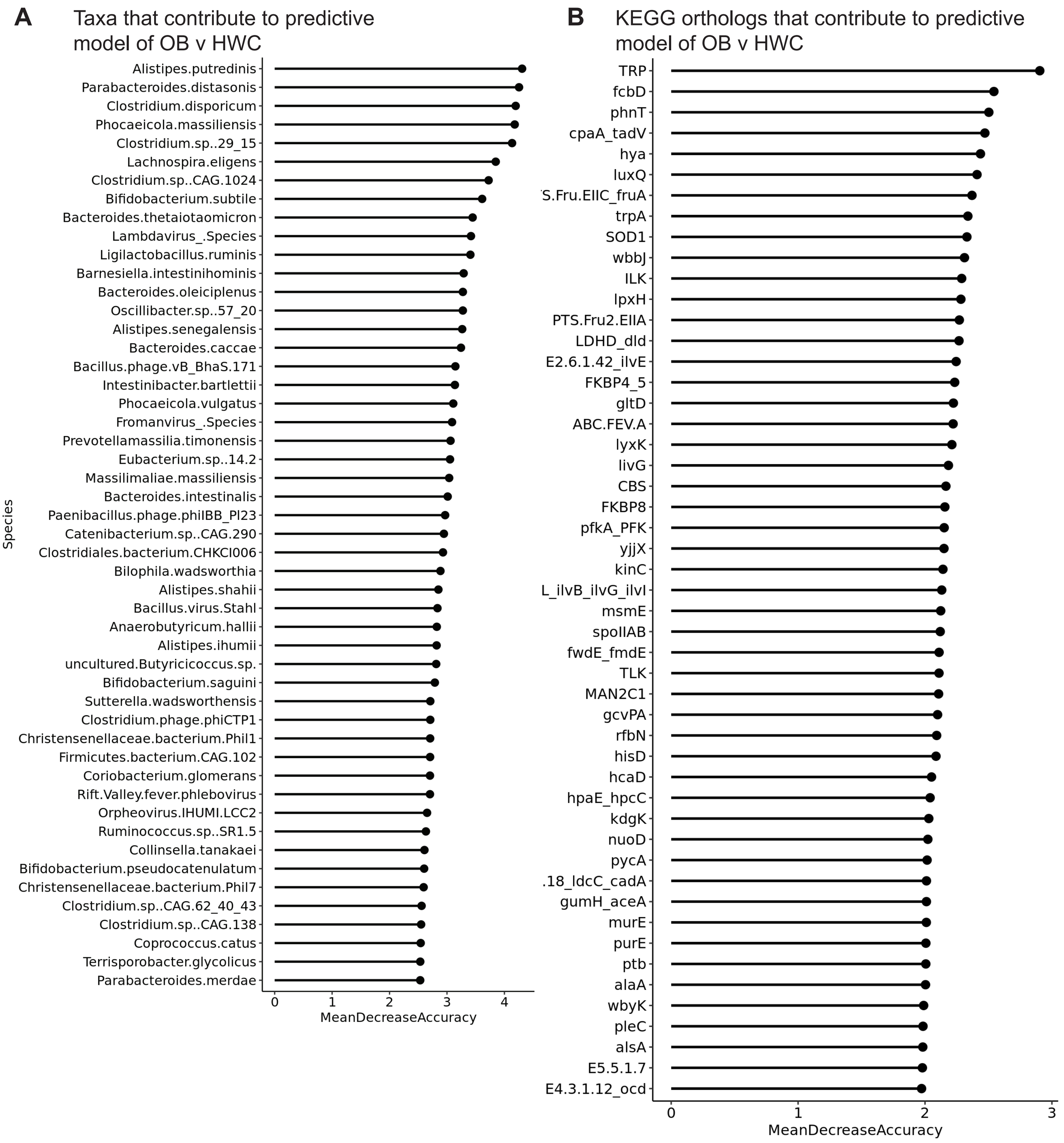


**Supplemental Figure 5. Random Forests machine learning analysis suggests (A) microbial taxa or (B) KEGG functional orthologs that contribute to model prediction of OB v HWC status**. "Mean decrease accuracy" score indicates the weight that each variable contributes to the accuracy of the predictive algorithm.

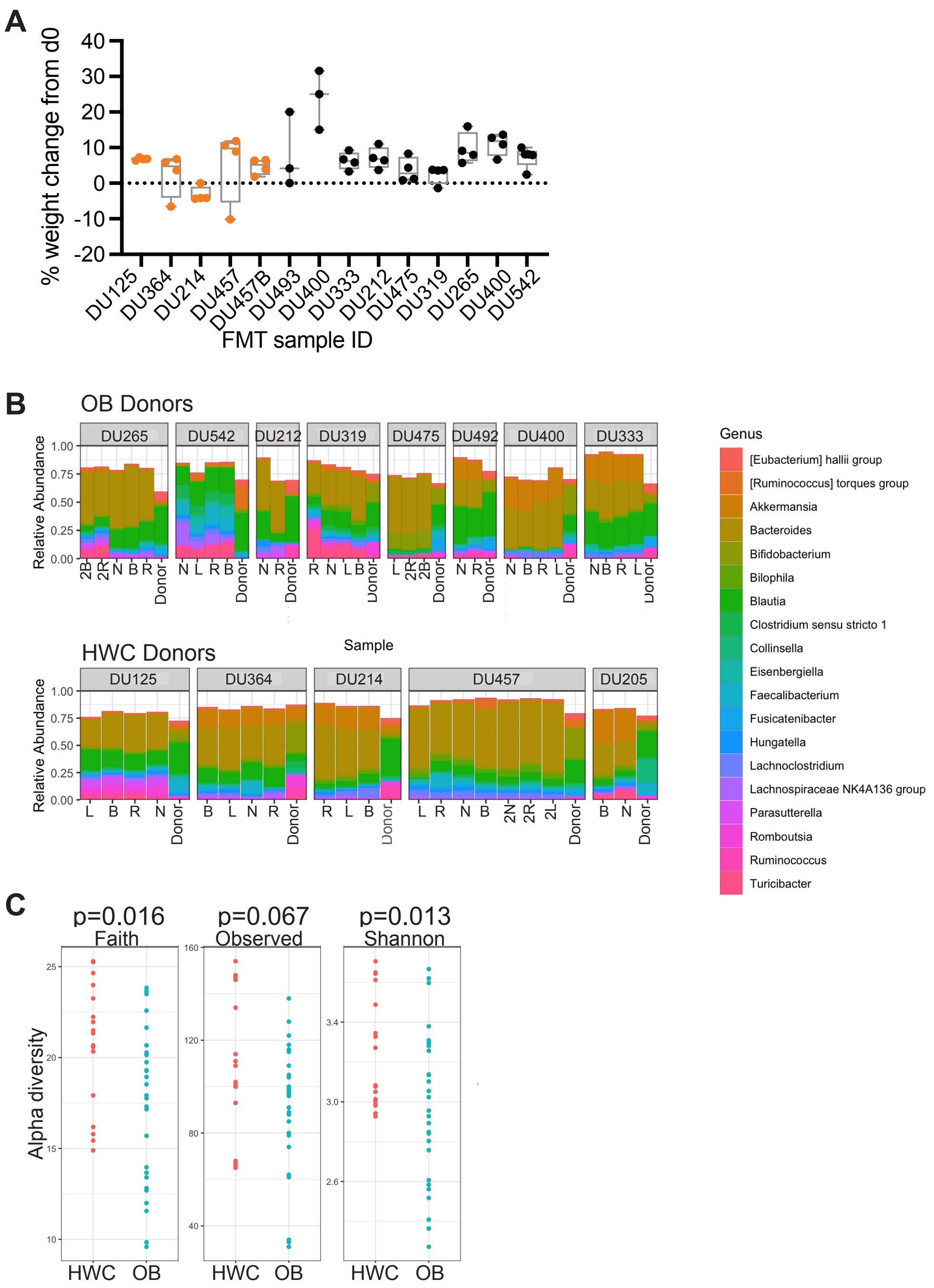
A
% weight change from d0
FMT sample ID
DU125
DU364
DU214
DU457
DU457B
DU493
DU400
DU333
DU212
DU475
DU319
DU265
DU400
DU542
B
OB Donors
DU265
DU542
DU212
DU319
DU475
DU492
DU400
DU333
Relative Abundance
Sample
HWC Donors
DU125
DU364
DU214
DU457
DU205
Donor
Genus
[Eubacterium] hallii group
[Ruminococcus] torques group
Akkermansia
Bacteroides
Bifidobacterium
Bilophila
Blautia
Clostridium sensu stricto 1
Collinsella
Eisenbergiella
Faecalibacterium
Fusicatenibacter
Hungatella
Lachnoclostridium
Lachnospiraceae NK4A136 group
Parasutterella
Romboutsia
Ruminococcus
Turicibacter
C
p=0.016
Faith
p=0.067
Observed
p=0.013
Shannon
Alpha diversity
HWC
OB

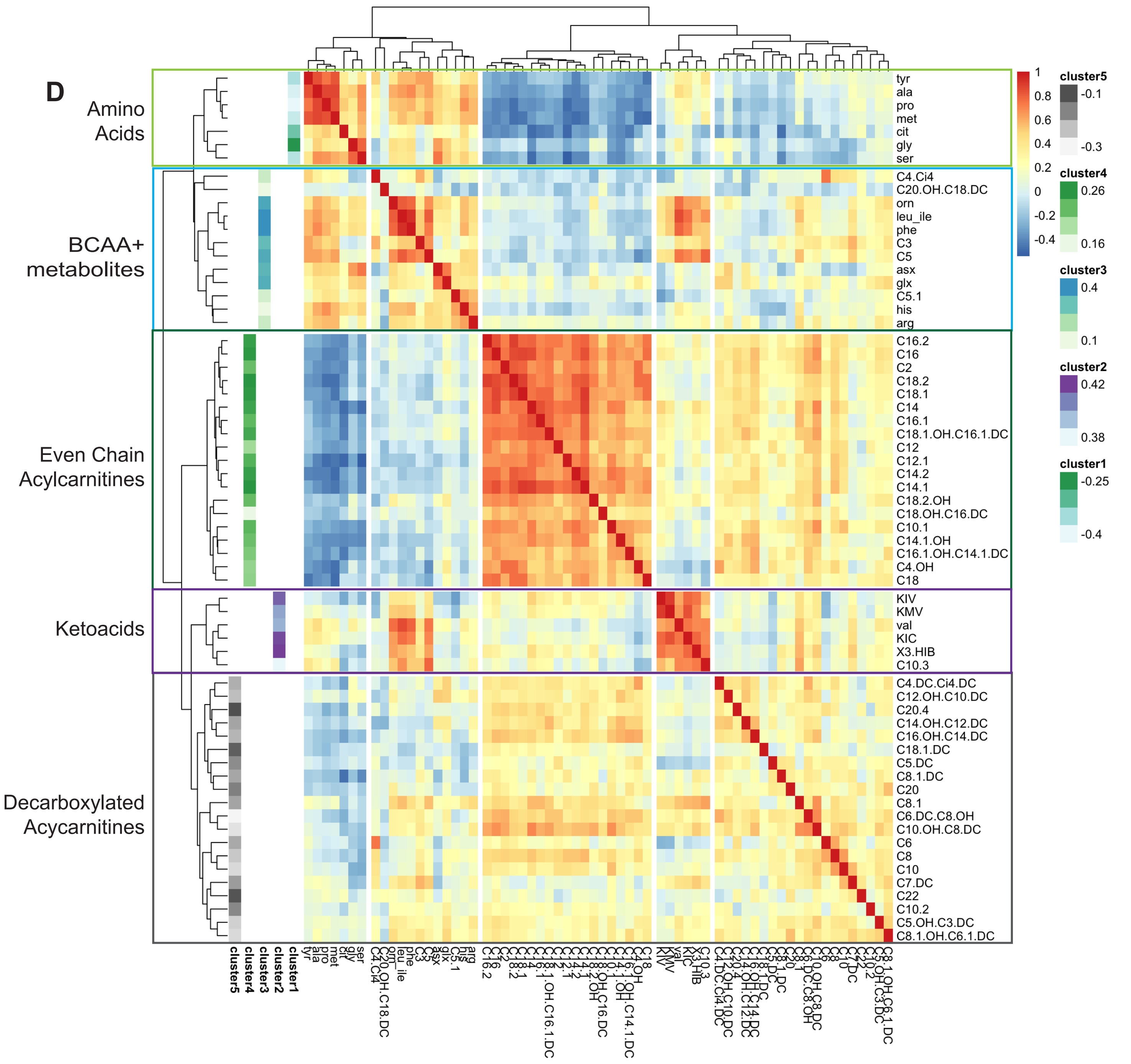


**Supplemental Figure 6. Donor status of fecal microbiome did not affect weight gain of recipient mice but did affect resulting fecal microbiome diversity. (A)** Percent weight change for germ free mice 2 weeks after receiving a fecal microbiome transplant from adolescents in the POMMS cohort. Each dot represents a single animal. Box plots represent mean and 1 standard deviation. Orange, HWC donor. Black, donors with OB. **(B)** Input gavage donor slurry (Donor) and mouse fecal microbiome community composition at 2 weeks post gavage as measured by 16S rRNA gene high throughput sequencing. R, N, L, B, 2R, 2R, 2N, 2B are individual animal tag labels. **(C)** Mouse fecal microbiome alpha diversity measures at 2 weeks post FMT gavage. **(D)** Results of hierarchical clustering of mouse serum metabolites for comparison with gut microbial taxa as referenced in main Figure 5G. Heatmap scale (red to blue) demonstrates intensity and direction of assocation, and Cluster color scales describe strength of association of each specific metabolite within the metabolite cluster.

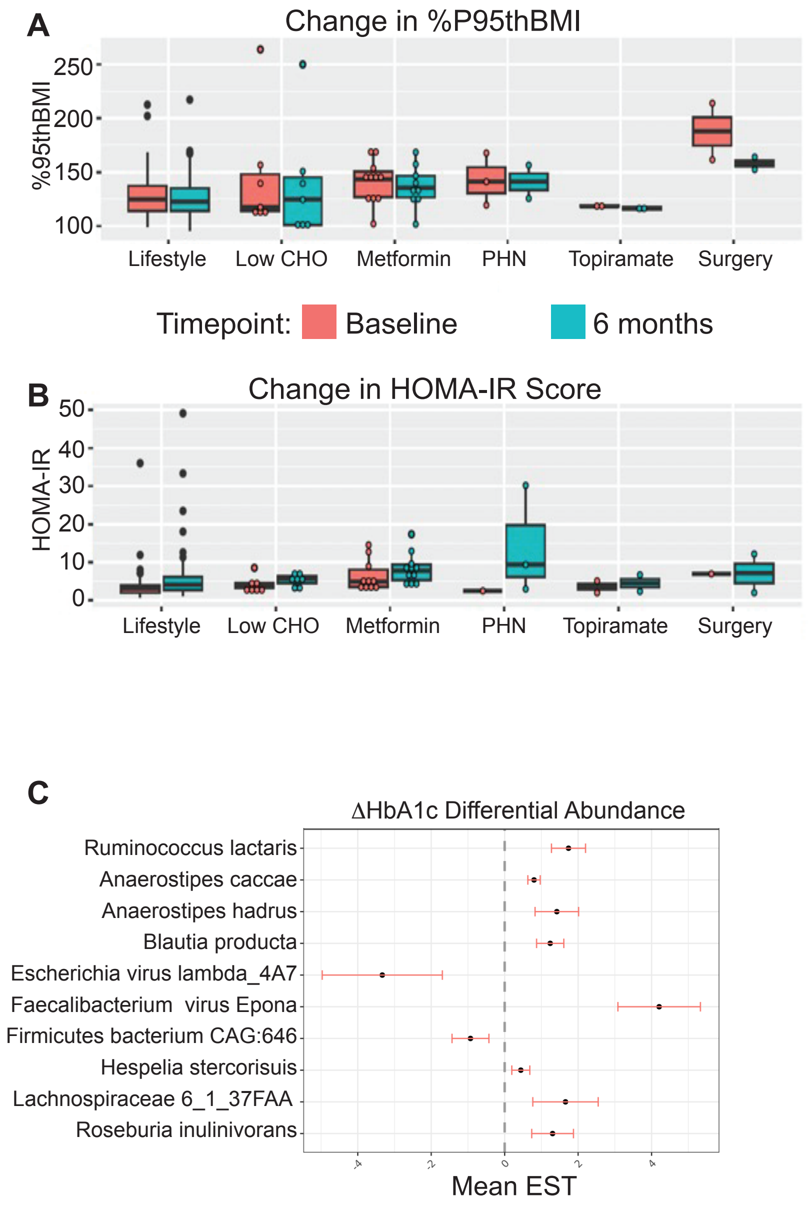

A
Change in %P95thBMI
%95thBMI
250
200
150
100
Lifestyle
Low CHO
Metformin
PHN
Topiramate
Surgery
Timepoint:
Baseline
6 months
B
Change in HOMA-IR Score
HOMA-IR
50
40
30
20
10
0
Lifestyle
Low CHO
Metformin
PHN
Topiramate
Surgery
C
ΔHbA1c Differential Abundance
Ruminococcus lactaris
Anaerostipes caccae
Anaerostipes hadrus
Blautia producta
Escherichia virus lambda_4A7
Faecalibacterium virus Epona
Firmicutes bacterium CAG:646
Hespelia stercorisuis
Lachnospiraceae 6_1_37FAA
Roseburia inulinivorans
Mean EST

**D**

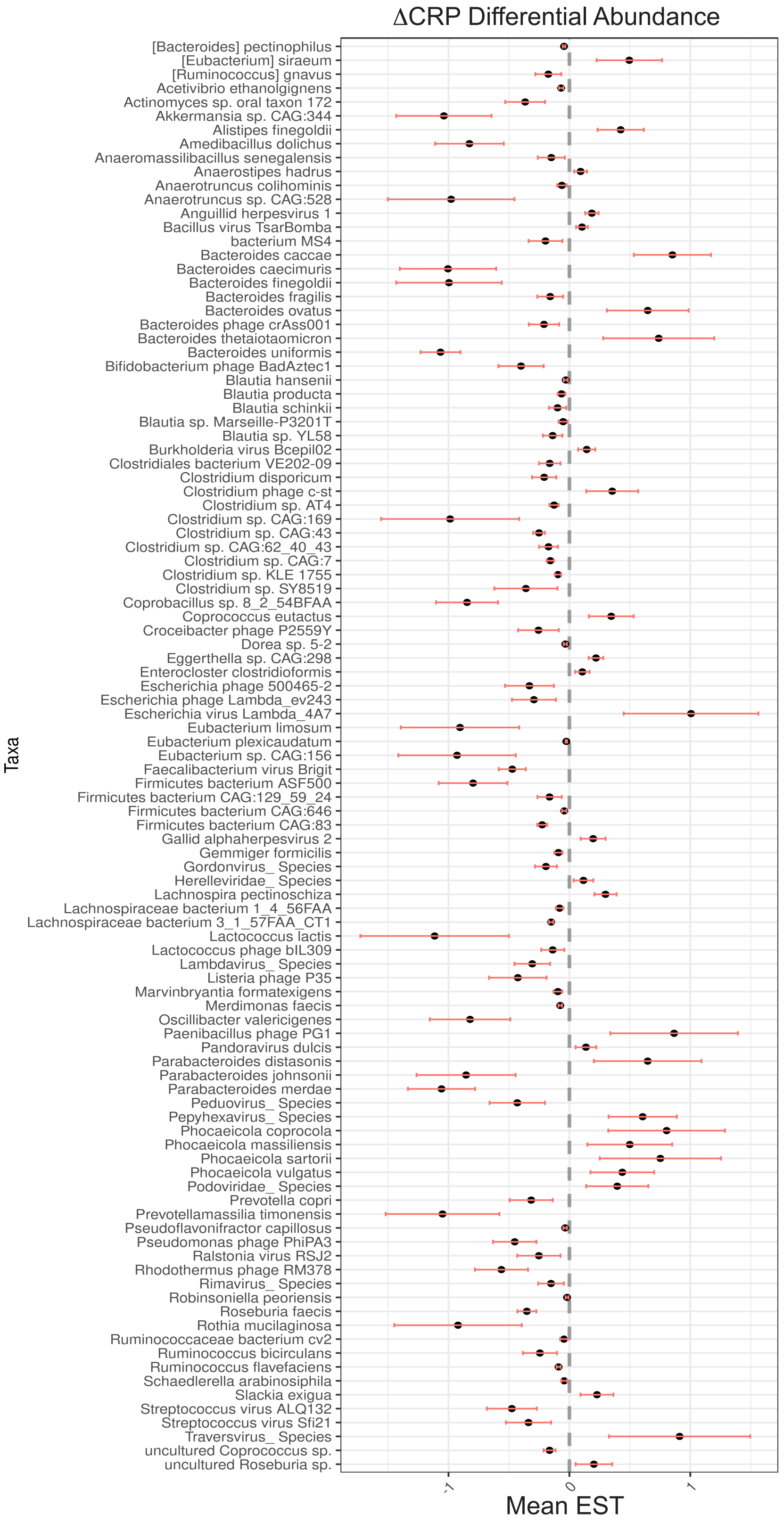


**Supplemental Figure 7. Most adolescents saw an increase in HOMA-IR over the observational period, and several baseline gut microbial taxa were significantly associated with changes in HbA1C and CRP in the OB cohort**. Mean and standard deviation of **(A)** %95thBMI and **(B)** HOMA-IR score at baseline (salmon) and 6 months (teal) for each treatment group within the observational trial. Baseline fecal microbiome taxa that were significantly associated with changes in **(C)** HbA1c or **(D)** C-reactive protein (CRP) levels over the observational period in the OB cohort. Abbreviations: CHO, carbohydrate; PHN, Phentermine.
